# Long-term ozone exposure associated cause-specific mortality risks with adjusted metrics by cohort studies: A systematic review and meta-analysis

**DOI:** 10.1101/2021.12.02.21267196

**Authors:** Haitong Zhe Sun, Pei Yu, Changxin Lan, Michelle Wan, Sebastian Hickman, Jayaprakash Murulitharan, Huizhong Shen, Le Yuan, Yuming Guo, Alexander T. Archibald

## Abstract

**BACKGROUND:** Long-term ozone (O_3_) exposure could lead to a series of non-communicable diseases and increase the mortality risks. However, cohort-based studies were still rather rare, and inconsistent exposure metrics might impair the credibility of epidemiological evidence synthetisation. To provide more accurate meta-estimation, this review updated the systematic review with inclusion of recent studies and summarised the quantitative associations between O_3_ exposure and cause-specific mortality risks based on unified exposure metrics.

**METHODS:** Research articles reporting relative risks between incremental long-term O_3_ exposure and causes of mortality covering all-cause, cardiovascular diseases, respiratory diseases, chronic obstructive pulmonary disease, pneumonia, ischaemic heart diseases, ischaemic stroke, congestive heart failure, cerebrovascular diseases, and lung cancer, estimated from cohort studies were identified through systematic searches in MEDLINE, Embase and Web of Science. Cross-metric conversion factors were estimated linearly by decadal of observations during 1990-2019. The Hunter-Schmidt random effect estimator was applied to pool the relative risks.

**RESULTS:** A total of 25 studies involving 226,453,067 participants (14 unique cohorts covering 99,855,611 participants) were included in the systematic review. After linearly adjusting the inconsistent O_3_ exposure metrics into congruity, the pooled relative risks (RR) associated with every 10 nmol mol^-1^ (ppbV) incremental O_3_ exposure, by mean of warm-season daily maximum 8-hour average metric, was: 1.014 with 95% confidence interval (CI) ranging 1.009–1.019 for all-cause mortality; 1.025 (95% CI: 1.010–1.040) for respiratory mortality; 1.056 (95% CI: 1.029–1.084) for COPD mortality; 1.019 (95% CI: 1.004–1.035) for cardiovascular mortality; and 1.096 (95% CI: 1.065–1.129) for congestive heart failure mortality. Insignificant mortality risk associations were found for ischaemic heart disease, cerebrovascular diseases and lung cancer.

**DISCUSSION:** This review covered up-to-date studies, expanded the O_3_-exposure associated mortality causes into wider range of categories, and firstly highlighted the issue of inconsistency in O_3_ exposure metrics. Non-intercept linear regression-based cross-metric RR conversion was another innovation, but limitation lay in the observation reliance, indicating further calibration with more credible observations available. Large uncertainties in the multi-study pooled RRs would inspire more future studies to corroborate or contradict the results from this review.

**CONCLUSION:** Adjustment for exposure metrics laid more solid foundation for multi-study meta-analysis, and wider coverage of surface O_3_ observations are anticipated to strengthen the cross-metric conversion in the future. Ever-growing numbers of epidemiological studies supported unneglectable cardiopulmonary hazards and all-cause mortality risks from long-term O_3_ exposure. However, evidences on long-term O_3_ exposure associated health effects were still scarce, and hence more relevant studies are encouraged to cover more population with regional diversity.

**REGISTRATION:** The review was registered in PROSPERO (CRD42021270637).

**FUNDING:** This study is mainly funded by UK Natural Environment Research Council, UK National Centre for Atmospheric Science, Australian Research Council and Australian National Health and Medical Research Council.

**Highlights:**

1. *Updated evidence for O*_*3*_*-mortality associations from 25 cohorts has been provided*.
2. *Adjusting various O*_*3*_ *exposure metrics can provide more accurate risk estimations*.
3. *Long-term O*_*3*_*-exposure was associated with increased mortality from all-causes, respiratory disease, COPD, cardiovascular disease and congestive heart failure*.

## 1 INTRODUCTION

Atmospheric ozone (O_3_) is a short-lived climate forcer.^1^ Besides warming the global atmosphere, O_3_ in the stratosphere can abate the radiation hazards from ultraviolet rays onto organisms, while O_3_ in the ambient air is of detrimental defects on ecosystem and human health,^2-4^ and hence health effects caused from exposure to surface O_3_ have become a serious public concern. Short-term (i.e. hours to days) exposure to high-level O_3_ can cause a series of acute symptoms like asthma, respiratory tract infection, myocardial infarction, and cardiac arrest;^5-8^ and long-term (i.e. over years) exposure can lead to chronic health outcomes including but not limited to preterm delivery, stroke, chronic obstructive pulmonary diseases, and cerebrovascular diseases.^9-12^ Long-term ambient O_3_ exposure was estimated to be responsible for over 0.36 million premature deaths globally in 2019 according to the Global Burden of Disease (GBD) report released by the Institute for Health Metrics and Evaluation (IHME).^13^

Systematic reviews summarising the associations between the adverse health outcomes, and both the short-term and long-term O_3_ exposures, have been performed in previous studies.^14-16^ Studies on short-term O_3_ exposure-induced morbidities are comparatively more abundant than the long-term O_3_ exposure studies where the epidemiological evidences are less congruous. Some deficiencies are spotted in the two reviews for long-term O_3_ exposure-associated mortality risk studies,^15, 16^ the primary issue of which is the inconsistent use of various O_3_ exposure metrics; however, no other reviews are found to remedy these flaws. As a secondary photolytic gaseous air pollutant, the warm-season and diurnal concentrations of surface O_3_ will be much higher than cool-season and nocturnal concentrations,^17, 18^ and thus the average and peak metrics of O_3_ concentrations shall be of drastically different realistic implications.^19^ Under this circumstance, directly pooling the relative risks scaled in different metrics might lead to biases.

Atkinson et al. (2016) explored 6 types of mortality causes, but searched the literatures only till 2015;^16^ while Huangfu et al. (2019) updated the searches to 2018, but only 3 types of mortality causes were considered.^15^ We thus determine to update the review on the health effects of O_3_ to include more categories of mortalities together with covering the most recent publications. Additionally, GBD estimations ascribed long-term O_3_-exposure induced all-cause mortality for chronic obstructive pulmonary disease,^13^ which might lead to underestimations without considering other causes. It is reasonable to deduce that long-term O_3_ exposure will exacerbate the mortality of certain diseases given that the short-term exposure increases the morbidity risks of the same diseases, and thus scrutinising epidemiological evidences for multiple causes of mortality will provide more credible supports to fill in this gap.

The primary innovation of our updated review is our taking full advantage of global systemic stationary observations to explore the feasibility of adjusting the various exposure metrics, and pooling the multi-study risks with the unified exposure metric, the mean of warm-season daily maximum 8-hour average, in response to the up-to-date suggestions from the Lancet global environmental health collaboration.^20^ Through this updated systematic review and meta-analysis on long-term O_3_ exposure associated cause-specific mortality risks, we aim to present and evaluate epidemiological evidences for 3 major questions not fully addressed by the previous 2 reviews, as (1) which mortality causes shall be ascribed to long-term O_3_ exposure; (2) have the risk associations changed given the latest studies with more mature research design and methodologies; and (3) how to estimate the quantities of the risk association strengths by the suggested exposure metric. Both our methods and discoveries are expected to inspire future O_3_-health studies, and support relevant policy-making to benefit the global population.

## 2. METHODS

### 2.1 Search strategy

We searched 3 research databases (MEDLINE, Embase, and Web of Science) from 1 September, 2015 till 1 February, 2022 to finish our systematic review and meta-analysis, updated from 2 previous reviews on long-term O_3_ exposure-associated mortality.^15, 16^ Search terms also referred to these 2 previous systematic reviews with modifications to enhance the inclusion of potential relevant studies, as we combined the keywords relevant to the cause-specific mortalities (i.e. “mortality”, “death”, “premature death”, “all-cause”, “non-accidental”, “cardiopulmonary”, “respiratory”, “chronic obstructive pulmonary disease”, “pneumonia”, cardiovascular, “lung cancer”, “cerebrovascular”, “stroke”, “ischaemic heart disease”, “congestive heart failure”), the pollutant of research interest (i.e. “ozone”), and qualified epidemiological study types (i.e. “long-term”, “cohort study”, “prospective”, “retrospective”, “longitudinal study”). The detailed search strategies were listed in Table S1. Health outcomes considered in the systematic review were: mortality from (1) all causes (AC, ICD9: 001-799, ICD10: A00-R99); (2) all respiratory diseases (RESP, ICD9: 460-519, ICD10: J00-J98); (3) chronic obstructive pulmonary diseases and allied conditions (COPD, ICD9: 490-496, ICD10: J19-J46); (4) all cardiovascular diseases (CVD, ICD9: 390-459, ICD10:I00-I98); (5) all cerebrovascular diseases (CEVD, ICD9: 430-438, ICD10: I60-I69); (6) ischaemic heart disease (IHD, ICD9: 410-414, ICD10: I20-I25); (7) congestive heart failure (CHF, ICD9: 428, ICD10: I50); (8) ischaemic stroke (ICD9: 434, ICD10: I61-I64); (9) pneumonia (ICD9: 480-487, ICD10: J12-J18); and (10) lung cancer (LC, ICD9: 162, ICD10: C33-C34).

### 2.2 Study eligibility criteria

As an updated systematic review, literatures identified in the previous 2 reviews underwent examination together with the newly retrieved ones. Studies were included during screening following the criteria as: (1) the epidemiological research should be conducted based on cohorts; (2) the exposure should include O_3_ as an individual risk factor; (3) the health outcomes should be all-cause or cause-specific deaths at individual level; (4) studies provided hazard ratio (HR), risk ratio (RR) or odds ratio (OR) and their 95% confidence intervals (CIs) clearly and reported by every increase unit (e.g. 10-ppbV) of exposure concentrations, assuming linear risk relationship with adjusting key confounders; (6) the study should be published as an original research article in scholarly peer-reviewed journals in English. For articles from the same cohort, only one single study covering the widest populations and the longest follow-up period was reserved for meta-analysis, unless the subgroups of participants and study follow-up periods are clearly stated to be of mild overlapping; We followed the Preferred Reporting Items for Systematic Review and Meta-Analyses (PRISMA) guidelines to process the included studies on ambient O_3_ exposure induced mortality.

### 2.3 Study selection and scrutinisation

All searched literatures were archived in Clarivate™ Analytics Endnote X9.3.1 reference manager software. Two literature review investigators (HZS and CL) conducted title and abstract pre-screening independently for all web-searched records and reviewed the full text for the pre-screened studies. Disagreements were resolved by discussions with a third reviewer (PY).

### 2.4 Data extraction

Details from each screened-out literatures were extracted and labelled for the purpose of meta-analysis, including (1) the authors with publication year as study labels of reference; (2) basic descriptive information of the study cohort embracing the cohort name, country, follow-up periods, numbers of cases and total participants, population genders and ethnics, exposure metrics, health outcomes, and major confounders; (3) the risk association effects preferably quantified in HR (and also RR/OR as substitute choices) per unit incremental exposure with 95% confidence interval (CI).

### 2.5 Study quality assessment

All screened-out studies underwent quality evaluation using the Quality Assessment Tool of Observational Cohort and Cross-Sectional Studies developed by National Institute of Health (NIH) (https://www.nhlbi.nih.gov/health-topics/study-quality-assessment-tools), aiming to ensure the studies considered for meta-analysis are adequately reliable. The assessments were cross-validated by two authors (HZS and CL) independently, with the third author (PY) supervising any disagreements. Table S2 listed 14 assessment items assigned with 1 score for each, and the tallied scores were translated into a literature-specific rating of quality. Studies scoring full-mark 14 were categorised to be “Good”, while 10-13 as “Fair” and <10 as “Poor”.

Besides applying the quality assessment tool to determine which reviewed studies should be included for meta-analysis, checking the epidemiological evidence quality from the included literatures for each cause of mortality was finished through the Grading of Recommendations Assessment, Development, and Evaluation (GRADE) system^21, 22^ to yield rating bands ranging across “high”, “moderate”, “low”, and “very low”. This grading system by default rated “high-quality” for cohort studies as the starting point of evaluation, and the rate would be downgraded by five limitations as the existence of (1) risk of bias examined by the Quality Assessment Tool (Table S2), (2) imprecision (i.e. studies did not report the central risk estimations with confidence intervals), (3) inconsistency (i.e. the directions of the estimated risks were controversial across studies), (4) indirectness (i.e. studies did not include the desired population, exposure, or health outcomes), and (5) publication bias (i.e. researchers tended to publish studies with positive results); and upgraded by three strengths as reporting (1) exposure-response trend, (2) residual confounding (i.e. adjusting the confounders highlighted the risks), and (3) strong associations. Publication biases were graphically presented by funnel plots,^23^ and statistically tested by trim-and-fill method.^24^ The review was registered in PROSPERO (CRD42021270637).

### 2.6 Exposure adjustment

#### 2.6.1 Unit unification

There were two major units used to quantify the surface O_3_ concentrations, nmol mol^-1^ or parts per billion by volume mixing ratio (ppbV) more frequently used by atmospheric modelling researchers,^17, 18, 25^ and milligram per cubic metre by mass concentration (µg/m^3^) widely used by public health studies.^12^ These two units are interchangeable to each other based on the ideal gas law *PV n RT*, if the air temperatures (T) and pressures (P) are given, as presented in eqs 1–4.

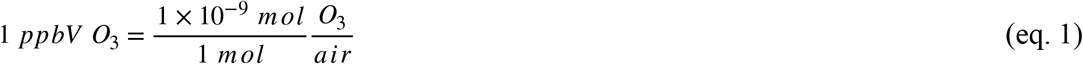

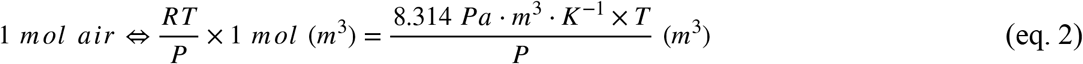

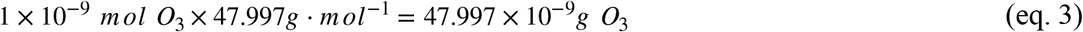

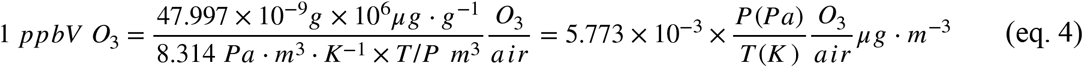

Assuming T = 298.65 K (25.5°C) and P = 101.325 kPa, the ppbV-µg/m^3^ conversion factor could be approximated as 1 ppbV ∼ 1.96 µg/m^3^. Though the surface air temperatures and pressures would vary across seasons, such simplification was still widely used in previous studies,^15, 26, 27^ being of more credibility for long-term surface O_3_ studies averaging the surface air temperatures and pressures at longer periods. For example, even at very low temperature of 270 K, the conversion factor was 2.17, which corroborated the stability of linear conversion.

#### 2.6.2 Metric unification

Surface O_3_, as a secondary photochemistry pollutant involving photolysis of NO_2_ to trigger chains of radical reactions, has concentrations that will vary significantly between day and night-time, and between warm and cool seasons, as discussed by numbers of studies.^17, 28-31^ Under this circumstance, various daily metrics to quantify the surface O_3_ concentrations emerged due to series of considerations, which however brought in more difficulties to assimilate epidemiological evidences. The previous reviews simply pooled the reported risk association strengths without adjusting the diverse metrics,^15, 16^ which we thought was a fatal defect requiring improvements.

We therefore designed to update the meta-analysis by unifying the exposure metrics for pooled O_3_ exposure-associated risks. As suggested by the U.S. EPA final report of *Air Quality Criteria for Ozone and Other Photochemical Oxidants*,^32^ linear relationships were assumed to estimate the cross-metric conversion factors using long-term reliable observations as the Tropospheric Ozone Assessment Report (TOAR) archive^19^ and China National Environmental Monitoring Centre (CNEMC, http://www.cnemc.cn/en/) in our review, and correlation matrix was used to validate that the presumptions of linearity were not violated. Both TOAR and CNEMC sites measured the surface O_3_ by means of the UV absorption technique with strict quality control so as to ensure the comparability of the records across different countries and regions.^33, 34^ We considered 6 complex metrics for mutual conversion as (1) annual mean of 24-hour daily average (ADA24), (2) 6-month warm season mean of 24-hour daily average (6mDA24), (3) annual mean of daily maximum 8-hour average (ADMA8), (4) 6-month warm season mean of daily maximum 8-hour average (6mDMA8), (5) annual mean of daily maximum 1-hour average (ADMA1), and (6) 6-month warm season mean of daily maximum 1-hour average (6mDMA1). Long-term averaging-based metric conversion could smooth the temporal variations resulting from the seasonal and geographical solar radiation variabilities. The linear conversion factors (*k*) were mathematically defined by eq 5, to adjust the original metric into the target one with irreducible regression errors *ε*.

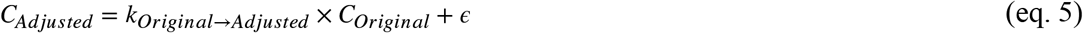

### 2.7 Meta-analysis

We collectively named relative risks (RR) for HR/RR/OR throughout our meta-analysis. All literature-reported RRs were converted into adjusted incremental risk ratios with a 10-ppbV O_3_-exposure increase by target metric (i.e. 6mDMA8 in this study), following eq 6 as shown below:

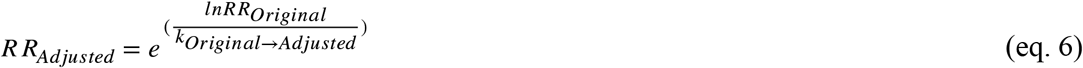

where *ln* is the natural logarithm, *RR*_*Original*_is the originally reported risk estimates scaled into 10-ppbV incremental exposure, and *k*_*Original*→*Adjusted*_ is the conversion factor for metric unification. Multi-study pooled risks with 95% confidence interval (CI) were calculated from the adjusted RRs by Hunter-Schmidt random effect meta-regression estimator to correct the potential errors and biases caused from the diversity of study population and methodologies.^35^

We applied the Higgins *I* ^2^to quantify the heterogeneity across studies. The Higgins statistics *I* ^2^ is defined as

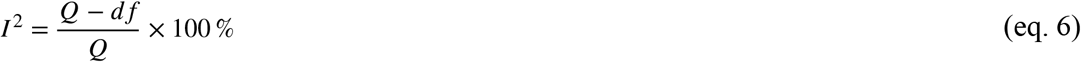

where Q is the Cochran’s non-parametric heterogeneity statistic assessing whether there are any cross-study differences in risks based on *x* ^2^ distribution and *df* is the corresponding degrees of freedom.^36^ Low *I* ^2^ values indicate no important heterogeneity observed and high *I* ^2^ values, especially >75%, indicate considerable heterogeneity.

Subgroup analyses were conducted by grouping the selected studies upon the gender, regions, O_3_ exposure metrics, and methodological reliability of individual exposure assignment; together with the adjustment of ethnicity, body mass index (BMI), smoking history, lifestyle features, and exposure to PM_2.5_ and NO_2_. Subgroups should contain at least 3 studies. Leave-one-out sensitivity analyses were also accomplished to test the robustness of synthesised overall risks by meta-analysis. All meta-analyses were performed in R 4.1.1 with packages *meta, metafor*, and *metainf*.

The most widely recognised approach to construct the integrated exposure-response (IER)^37^ relationships required sufficient epidemiological studies to comprehensively sample the population exposure levels. However, studies on long-term O_3_ exposure health effects were relatively limited, under which circumstance we made methodological modifications to make better use of the variabilities in exposure levels by statistically imputing the exposure distributions for each study from the provided statistics (e.g. mean, standard deviation, and percentiles) for curve fitting as elaborated in Supplementary Text S1. Supplementary Text S2 described the detailed procedures of exposure distribution imputations with a demonstration provided in S3, through which high uncertainties were still observed in the fitted IER curves due to insufficient epidemiological evidences.

## 3. RESULTS

### 3.1 Study characters

From the 3 databases during September 2015 till February 2022, a total of 339 studies (77 from MEDLINE, 102 from Embase, and 160 from Web of Science) were searched; and together with 34 additional literatures added manually from the 2 previous systematic reviews,^15, 16^ 373 studies underwent duplication censoring, deleting 101 duplicated studies. After detailed scrutinisation for 272 studies, a total of 25 studies concerning long-term O_3_ exposure and multi-cause mortalities were finally enrolled for quality evaluation, meta-analysis and further discussions (**Figure 1**).^38-62^ **Table 1** summarised the basic information of the 25 included studies sorted by the publication year and surname of the first author.

**Table 1.** Summary of cohort characteristics included for meta-analysis.

| Study | Cohort | Country | Follow-up Duration | Population Type | Sample Size | Sex | Age | Key Confounding Adjustment | Mortality Causes |
| --- | --- | --- | --- | --- | --- | --- | --- | --- | --- |
| Abbey et al. 1999<sup>38</sup> | AHS | USA | 1977-1992 | Occupational | 6,182 | FM | 27-95 | age, sex, BMI, smoking, individual demographic features , lifestyle features<sup>+</sup>, medical history | AC, RESP, LC |
| Lipfert et al. 2006<sup>39</sup> | WU-EPRI | USA | 1976-1996 | General | 67,108 | M | 51 (12)<sup>§</sup> | age, ethnicity, BMI, smoking, traffic density, NO<sub>2</sub>, CO | AC |
| Jerrett et al. 2009<sup>40</sup> | ACS CPS II | USA | 1977-2000 | General | 448,850 | FM | ≥ 30 | age, sex, ethnicity, BMI, smoking, individual demographic features, lifestyle features, PM<sub>2.5</sub> | AC, RESP, CVD, IHD |
| Krewski et al. 2009<sup>41</sup> | ACS CPS II | USA | 1982-2000 | General | 488,370 | FM |  |  | AC, IHD, LC |
| Smith et al. 2009<sup>42</sup> | ACS CPS II | USA | 1982-2000 | General | 352,242 | FM |  |  | AC, RESP, CVD |
| Lipsett et al. 2011<sup>43</sup> | CTS | USA | 1998-2005 | Occupational | 124,614 | F | ≥ 20 | age, ethnicity, BMI, smoking, lifestyle features, medical treatment | AC, RESP, CVD, IHD, CEVD, LC |
| Zanobetti et al. 2011<sup>44</sup> | Medicare | USA | 1985-2006 | General | 8,894,473 | FM | ≥ 65 | age, sex, ethnicity, medical history | COPD, CHF |
| Carey et al. 2013<sup>45</sup> | CPRD | UK | 2003-2007 | General | 824,654 | FM | 40-89 | age, sex, BMI, smoking, individual demographic features | AC, RESP, LC |
| Jerrett et al. 2013<sup>46</sup> | ACS CPS II | USA | 1982-2000 | General | 73,711 | FM | 57 (11) | age, sex, smoking, individual demographic features, lifestyle features | AC, RESP, CVD, IHD, LC |
| Bentayeb et al. 2015<sup>47</sup> | GAZEL | France | 1989-2013 | Occupational | 20,327 | FM | 44 (4) | age, sex, BMI, smoking, individual demographic features, lifestyle features | AC, RESP, CVD |
| Crouse et al. 2015<sup>48</sup> | CANCHEC | Canada | 1991-2006 | General | 2,521,525 | FM | ≥ 25 | age, sex, individual and area-level demographic features, PM<sub>2.5</sub>, NO<sub>2</sub> | AC, RESP, COPD, CVD, IHD, CEVD, LC |
| Tonne et al. 2016<sup>49</sup> | MINAP | UK | 2003-2010 | MI Survivors<sup>*</sup> | 18,138 | FM | 68 (14) | age, sex, ethnicity, smoking, medical history, area-level demographic features | AC |
| Turner et al. 2016<sup>50</sup> | ACS CPS II | USA | 1982-2004 | General | 669,046 | FM | ≥ 30 | age, sex, BMI, smoking, individual and area-level demographic features, PM<sub>2.5</sub>, NO<sub>2</sub> | AC, RESP, COPD, CVD, CHF, IHD, CEVD |
| Di et al. 2017<sup>51</sup> | Medicare | USA | 2000-2012 | General | 60,925,443 | FM | ≥ 65 | age, sex, ethnicity, BMI, smoking, individual and area-level demographic features, meteorological features, PM<sub>2.5</sub> | AC |
| Weichenthal et al. 2017<sup>52</sup> | CANCHEC | Canada | 2001-2011 | General | 2,448,500 | FM | 25-89 | age, sex, ethnicity, individual and area-level demographic features | AC, RESP, CVD |
| Cakmak et al. 2018<sup>53</sup> | CANCHEC | Canada | 1991-2011 | General | 2,291,250 | FM | ≥ 25 | age, sex, individual demographic features, PM<sub>2.5</sub> | AC, COPD, IHD, LC |
| Hvidtfeldt et al. 2019<sup>54</sup> | DDCH | Denmark | 1993-1997 | General | 49,596 | FM | 50-64 | age, sex, BMI, smoking, individual and area-level demographic features, noise | AC, RESP, CVD |
| Kazemiparkouhi et al. 2019<sup>55</sup> | Medicare | USA | 2000-2008 | General | 22,159,190 | FM | ≥ 65 | age, sex, ethnicity, area-level demographic features, PM<sub>2.5</sub> | AC, RESP, COPD, CVD, IHD, CHF, CEVD, LC |
| Lim et al. 2019<sup>56</sup> | NIH-AARP | USA | 1995-2011 | General | 548,780 | FM | 50-71 | age, sex, ethnicity, BMI, smoking, individual demographic features, PM<sub>2.5</sub>, NO<sub>2</sub>, daily maximum temperature | AC, RESP, COPD, CVD, IHD, CHF, CEVD, LC |
| Paul et al. 2020<sup>57</sup> | ONPHEC | Canada | 1996-2015 | Diabetes | 452,590 | FM | 35-85 | age, sex, area-level demographic features | CVD |
| Shi et al. 2021<sup>58</sup> | Medicare | USA | 2001-2017 | General | 44,684,756 | FM | ≥ 65 | age, sex, ethnicity, BMI, smoking, individual and area-level demographic features, lifestyle features, PM<sub>2.5</sub>, NO<sub>2</sub>, medical history | AC |
| Strak et al. 2021<sup>59</sup> | ELAPSE | 6 countries<sup>†</sup> | 1985-2015 | General | 325,367 | FM | 49 (13) | age, sex, ethnicity, BMI, smoking, individual and area-level demographic features, PM<sub>2.5</sub>, NO<sub>2</sub>, BC | AC, RESP, COPD, CVD, IHD, CEVD |
| Yazdi et al. 2021<sup>60</sup> | Medicare | USA | 2000-2016 | General | 44,430,747 | FM | ≥ 65 | age, sex, ethnicity, BMI, smoking, individual and area-level demographic features, lifestyle features, PM<sub>2.5</sub>, NO<sub>2</sub>, medical history | AC |
| Bauwelinck et al. 2022<sup>61</sup> | BC2001 | Belgium | 2001-2011 | General | 5,474,470 | FM | ≥ 30 | age, sex, individual and area-level demographic features, PM<sub>2.5</sub>, NO<sub>2</sub> | AC, RESP, CVD, LC |
| Stafoggia et al. 2022<sup>62</sup> | ELAPSE | 7 countries<sup>‡</sup> | 2000-2017 | General | 28,153,138 | FM | ≥ 30 | age, sex, ethnicity, BMI, smoking, individual and area-level demographic features, PM<sub>2.5</sub>, NO<sub>2</sub>, BC | AC, RESP, CVD, LC |

Cohort abbreviations: AHSMOG, *Adventist Health Study of Smog*; WU-EPRI, *Washington University–Electric Power Research Institute*; ACS CPS, *American Cancer Society Cancer Prevention Study*; CTS, *California Teacher Study*; CPRD, *Clinical Practice Research Datalink*; GAZEL, *GAZ de France and Électricité*; CANCHEC, *Canadian Census Health and Environment Cohort*; MINAP, *National Audit of Myocardial Infarction Project*; DDCH, *Danish Diet, Cancer and Health*; NIH-AARP, *National Institute of Health, American Association of Retired Persons*; ONPHEC, *Ontario Population Health and Environment Cohort*; BC2001, *Belgian 2001 Census*.
<sup>¶</sup> Demographic features included marital status, education attainment, employment status and occupational class, aboriginal ancestry, visible minority ethnicities, immigrant status and residence location (urban or rural), income level and socioeconomic status (SES), and regional population density. Different studies adjusted various combinations of demographic features.
<sup>⊥</sup> Lifestyle features included consumptions of alcohol, dietary fat, vegetables, fruits (dietary fibre), and vitamins, together with physical activity frequency. Different studies adjusted various combinations of lifestyle features.
<sup>§</sup> Population ages were reported by mean with standard deviation (in bracket).
<sup>\*</sup> MI, Myocardial Infarction.
<sup>†</sup> Sweden, Denmark, France, Netherland, Germany and Austria.
<sup>‡</sup> Belgium, Denmark, England, Netherland, Norway, Switzerland and Italy.

**Figure 1.**
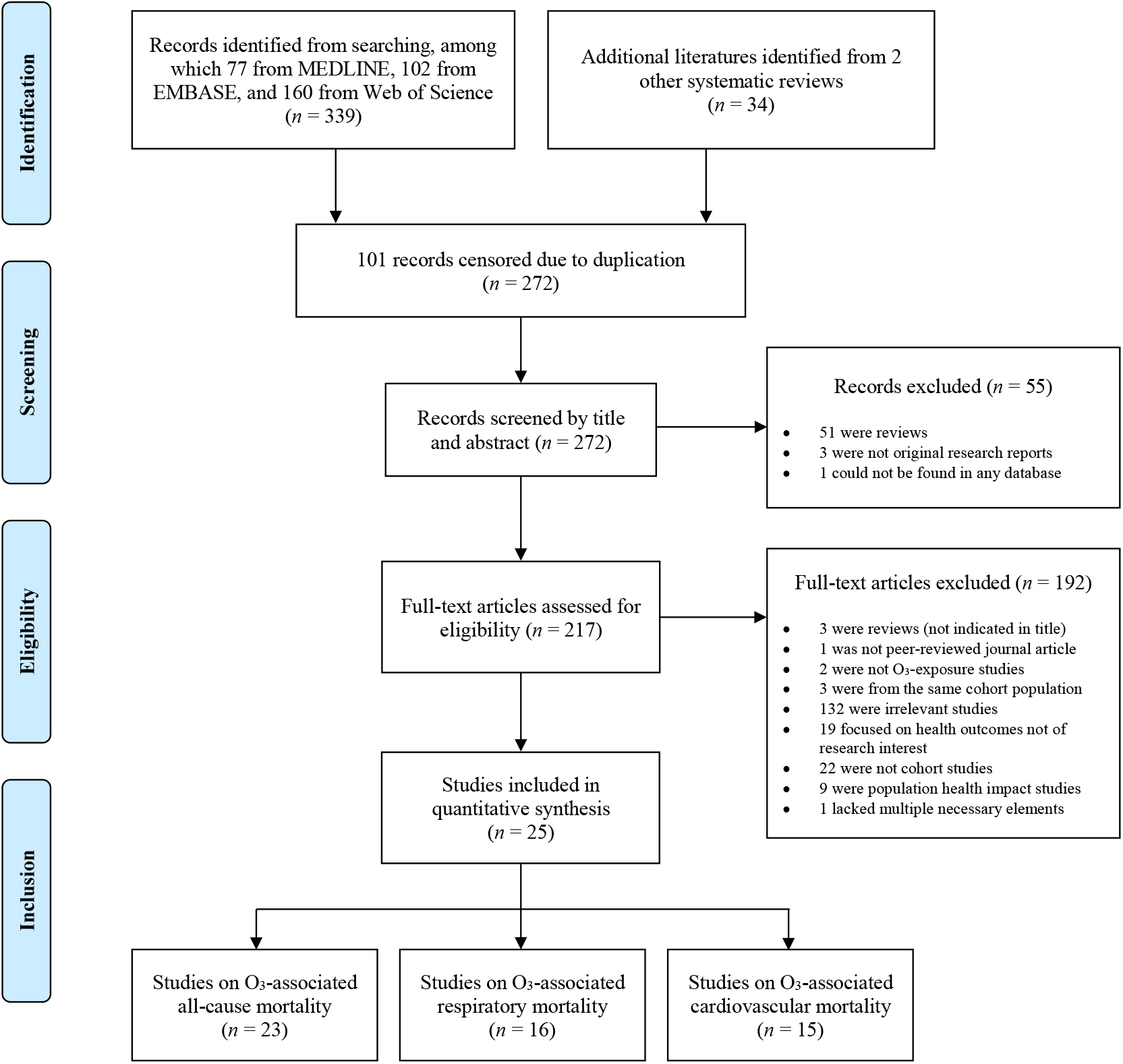
Schematic flowchart of study assessment and selection processes for literature review and meta-analysis.

### 3.2 Metrics and exposure assignments

Our updated systematic review stressed more on the exposure metrics and methodologies to obtain O_3_ exposure, as summarised in **Table 2**. Abbey et al. (1999),^38^ Jerrett et al. (2013)^46^ and Lipsett et al. (2011)^43^ did not state the metric they used clearly, but based on comparisons between the reported surface O_3_ concentrations and TOAR observational archives, we reasonably assumed ADA24 for the first study, and ADMA8 for the rest two. Details of the metric matching were given in Supplementary Text (S4). Lipfert et al. (2006)^39^ used the highest 95^th^ percentile by hourly resolved O_3_ concentrations as the peak exposure metric, which was only used in this one singular study, and hence approximated to DMA1. Krewski et al. (2009)^41^ and Smith et al. (2009)^42^ were both studies on ACS CPS II, and thus the same exposure assignment methodologies and metrics were assumed as Jerrett et al. (2009).^40^ Likewise, Cakmak et al. (2018)^53^ and Weichenthal et al. (2017)^52^ were assumed to inherit Crouse et al. (2015)^48^ as all these 3 studies were on CANCHEC. Warm season was defined as 6 months from April to September in terms of the northern hemisphere by default, but we made no exceptions to 3 studies as Zanobetti et al. (2011)^44^ using May to September, and Crouse et al. (2015)^48^ and Paul et al. (2020)^57^ using May to October, due to limited number of studies searched.

**Table 2.**
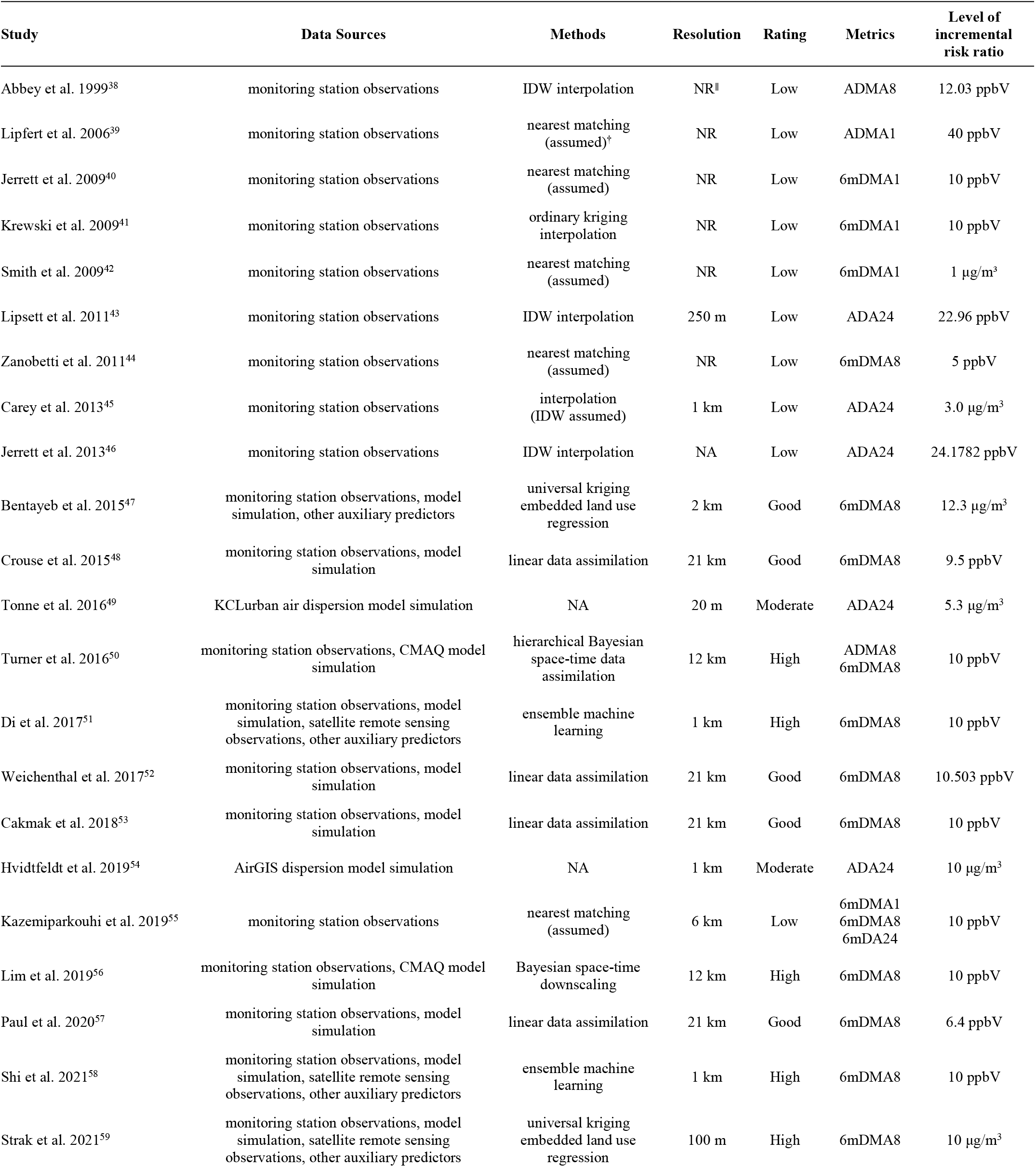
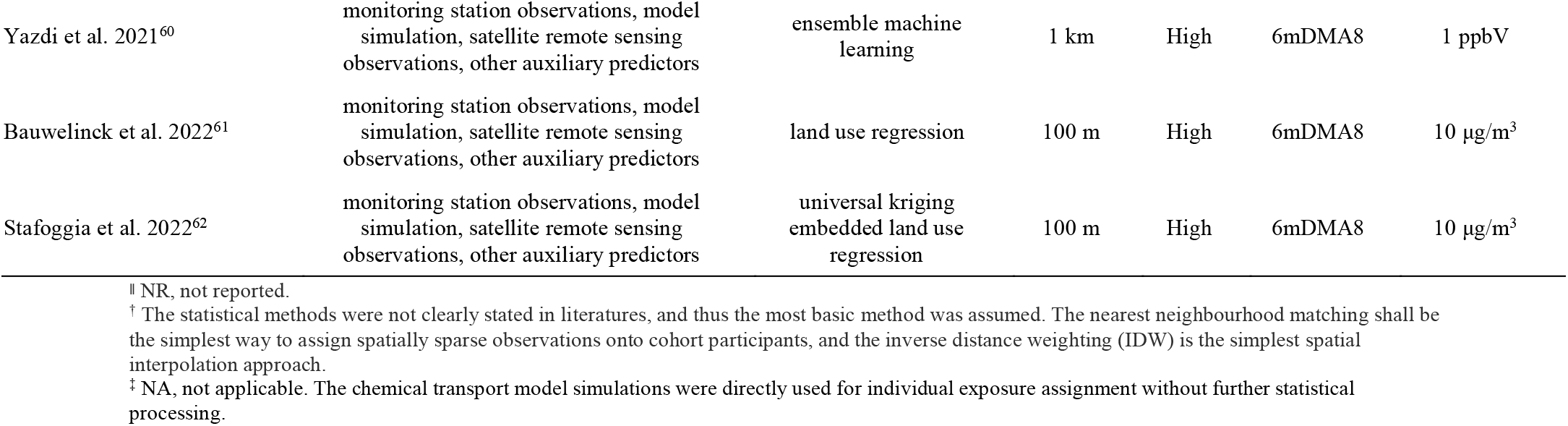
Data sources and statistical methods of O_3_ exposure assignment. Methodological ratings were based on spatial interpolation and multi-data assimilation approaches. Spatial resolutions, exposure metrics, and levels of incremental risk ratio were also listed.

Across all included studies, multiple methods were applied to obtain gap-free surface O_3_ concentrations for individual-level exposure assignment. The most basic way was the nearest neighbour matching between participant residential locations and *in situ* observation sites, which were more frequently used in earlier studies.^39, 40^ A comparatively more complicated way was statistical spatial interpolation, by inverse distance weighting^46^ or ordinary kriging^41^. Full spatial coverage products, such as satellite-based remote-sensing^51^ and chemistry transport models^56^, were used in some studies by supervised-learning-based data fusion techniques including but not limited to universal kriging embedded land use regression,^47^ Bayesian hierarchical model^50^ and ensemble learning^51^ to enhance the spatial extrapolation accuracy, which were evaluated to be of higher credibility than the basic ones described previously. All basic interpolation methods using merely the observations were rated as “Low”; applying chemical transport model simulations without calibration from the observations as “Moderate”; linearly coupling the observations with simulations as “Good”; and multi-source data assimilation by means of more sophisticated approaches as “High”. To sum up, 8 studies were rated “High”, 5 were “Good”, 2 were “Moderate”, and 10 were “Low”. Methodological progresses with time were evident as manifested in **Table 2**, prefiguring an explosion of population-based environmental health studies in the age of big data.

Based on the TOAR and CNEMC *in situ* observations, the cross-metric non-intercept linear conversion factors were estimated with regression accuracies given in **Figure 2**. Synthesised from the recent relevant studies, the 6mDMA8 metric was more recommended to highlight the peak exposure; and therefore, we chose to convert the originally reported RRs uniformly into the 6mDMA8 scale as standard. The O_3_ exposure levels by the original and unified metric were listed in Supplementary Text S1. Demonstrations for the conversion interpretation and procedures were presented in Supplementary Text S5, respectively.

**Figure 2.**
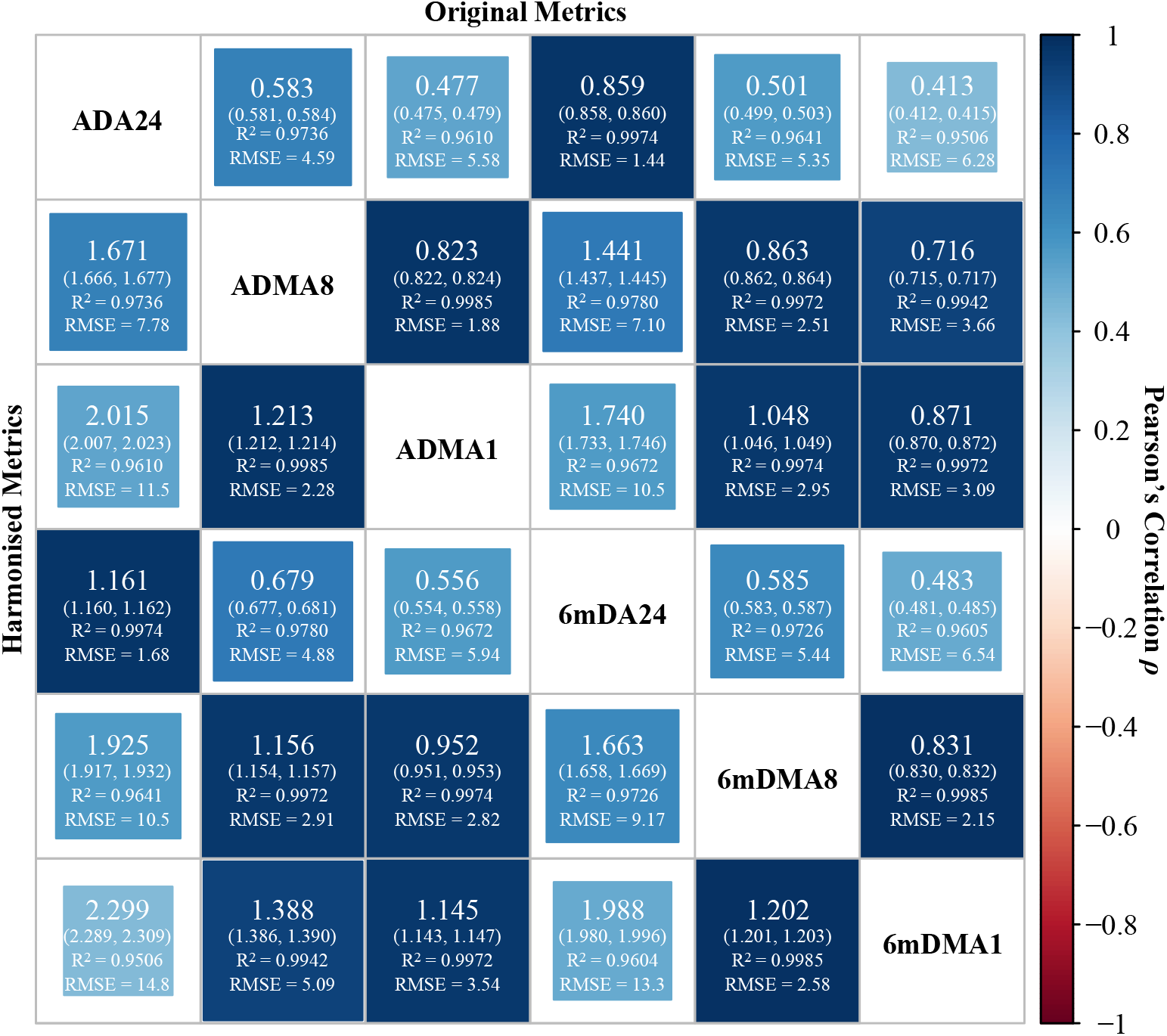
Cross-metric linear relationships and conversion accuracies. The cross-metric linear relationships were scaled by Pearson’s correlation coefficients. The cross-metric conversion factors with 95% confidence intervals (95% CI) were estimated by non-intercept linear regression models, accompanied with fitting accuracies quantified by coefficient of determination (R^2^) and root mean square error (RMSE) in ppbV. The conversion factors were defined as multiples from the original metric by column into the target harmonised metric by row, e.g. ADMA8 = 1.671 ADA24, R^2^ = 0.9736, RMSE = 7.78 ppbV. Note that by non-intercept linear regression, the values of R^2^ should no longer be equal to the squared Pearson’s linear correlation coefficients. As the cross-metric conversion coefficients were estimated statistically, indirect conversions were not recommended, since regression noises restricted the validity of equation *k*_*A→B*_ = *k*_*A→C*._ *k*_*C→B*_.

### 3.3 Meta-analysis results

We conducted meta-analyses for long-term O_3_ exposure-associated into 10 categories of mortalities as (1) all causes (AC), (2) all respiratory diseases (RESP), (3) chronic obstructive pulmonary diseases and allied conditions (COPD), (4) all cardiovascular diseases (CVD), (5) all cerebrovascular diseases (CEVD), (6) ischaemic heart disease (IHD), (7) congestive heart failure (CHF), and (8) lung cancer (LC), with the exposure metrics adjusted into 6mDMA8.

#### 3.3.1 All-cause mortality

A total of 23 studies were included into O_3_ exposure-associated all-cause mortality meta-analysis, pooling the overall risk into RR = 1.014 (95% CI: 1.009–1.019, I^2^: 97.8%) with every 10-ppbV incremental exposure by 6mDMA8 as presented in **Figure 3**. Sub-group meta-analysis by originally reported metrics concluded the significances of risks vary across metrics, as high-concentration highlighted metrics like 6mDMA8 were of the highest positive risk (RR = 1.022, 95% CI: 1.014–1.031) while the smoothed metric ADA24 reported negative association (RR = 0.980, 95% CI: 0.960–1.001), as shown in Figure S1. Grouped by study regions, significant discrepancies of the risk pattern was found (Figure S2), as the studies in North America revealed positive associations as RR = 1.019 (95% CI: 1.014–1.024), while researches on European populations showed reversed risks as RR = 0.910 (95% CI: 0.827–1.001), though not significant. The cross-region divergences did not necessarily indicate differences in population vulnerability, as (1) less and younger study population, (2) shorter follow-up durations, and (3) use of smoothed exposure metrics for studies in Europe could all potentially obscure the potential risk associations. Subgroup analysis manifested that high inter-study heterogeneities originated from metric inconsistency, methodological reliability of individual exposure assignment, and population variabilities, as encapsulated in Table S3. The funnel plot was visually symmetrical (Figure S3), and studies reporting risks below the pooled value were even slightly more, indicating no detected severe potential publication biases.

**Figure 3.**
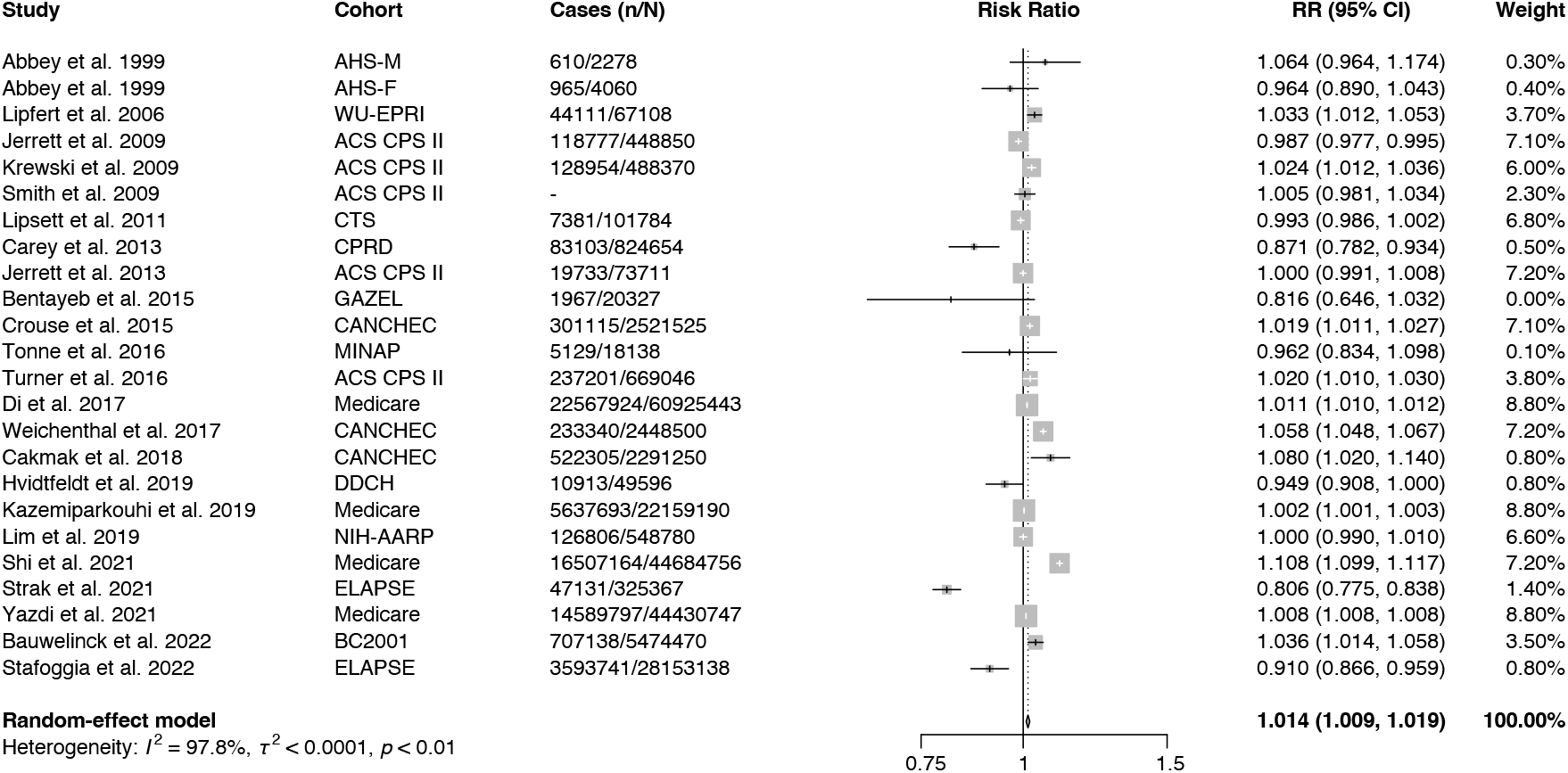
Pooled estimates of all-cause mortality risk associated with every 10-ppbV incremental O_3_ exposure by 6mDMA8 metric. Size of the shaded squares in the forest plot represents the weight of each study estimated by random-effect model.

No significant inter-gender differences were observed based on the limited studies reporting gender-specific risk association strengths. Further subgroup analyses were unfeasible due to the lack of reporting in the literature. Alternatively, grouped RRs were estimated based on whether the original researches had adjusted the confounding effects from ethnicity, body mass index (BMI), smoking history, lifestyle features, exposure levels of PM_2.5_ and NO_2_, and no inter-group divergences were observed (Table S3).

#### 3.3.2 Respiratory mortality

Meta-analysis for O_3_ exposure-associated all respiratory mortality includes 16 studies, pooling which gave the overall RR = 1.025 (95% CI: 1.010–1.040, I^2^: 83.9%) for every 10-ppbV incremental O_3_ exposure by 6mDMA8 (**Figure 4**). Based on sub-group meta-analysis for different metrics (Figure S4), peak metrics showed more significant increasing risks than ADA24, where most of the heterogeneities were from (I^2^ = 87.0%). Cross-metric divergences were generally higher than intra-metric discrepancies. Studies on North America populations showed better homogeneity in positive risks (RR = 1.029, 95% CI: 1.011–1.047, I^2^ = 71.1%, Figure S5) than the European cohorts, pooling from which the overall risks were congruously insignificant (RR = 0.941, 95% CI: 0.856– 1.036, I^2^ = 91.2%). For O_3_-COPD mortality association, the pooled RR was 1.056 (95% CI: 1.029–1.084, I^2^ = 94.5%) for 10-ppbV incremental O_3_ exposure by 6mDMA8 from 7 studies. No apparent positive publication biases were detected for both respiratory and COPD mortalities from the funnel plot (Figure S3).

**Figure 4.**
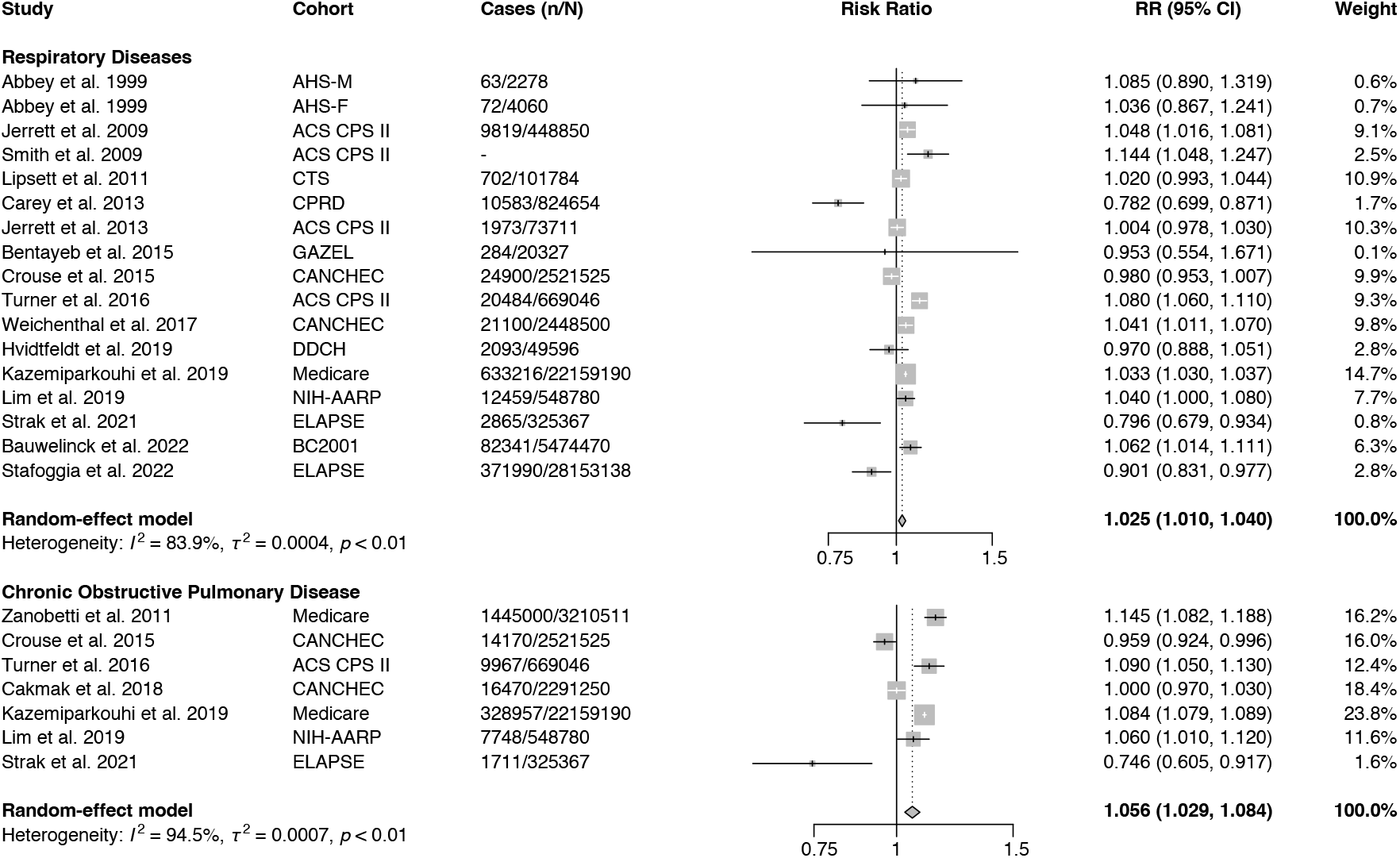
Pooled estimates of respiratory diseases and COPD mortality risks associated with every 10-ppbV incremental O_3_ exposure by 6mDMA8 metric.

#### 3.3.3 Cardiovascular mortality

A total of 15 studies were included to pool the overall O_3_ exposure-induced CVD mortality risks as RR = 1.019 (95% CI: 1.004–1.035, I^2^ = 97.7%) for each 10-ppbV additional O_3_ exposure by 6mDMA8 (**Figure 5**). Given the fact that the lower bound of uncertainty interval was so close to the null hypothesis (i.e. RR = 1), the positive risk association found in this review could be controversial, and thus would require more studies to support or refute the finding. Heterogeneities (I^2^ > 79.2%) were observed through all 3 metric-grouped studies as presented in Figure S6. Positive risk associations were found on 10 North American cohorts (RR = 1.036, 95% CI: 1.019–1.053) while oppositely for 5 European cohorts (RR = 0.934, 95% CI: 0.865–1.008), as shown in Figure S7. There was no need to be concerned with the publication bias, and no more inter-group divergences were spotted except for grouping by exposure assignment methodological credibility (Table S3). The pooled risk for the congestive heart failure-induced mortality from 4 studies was RR = 1.074 (95% CI: 1.054–1.093, I^2^ = 85.8%) with every 10-ppbV incremental O_3_ exposure by 6mDMA8.

**Figure 5.**
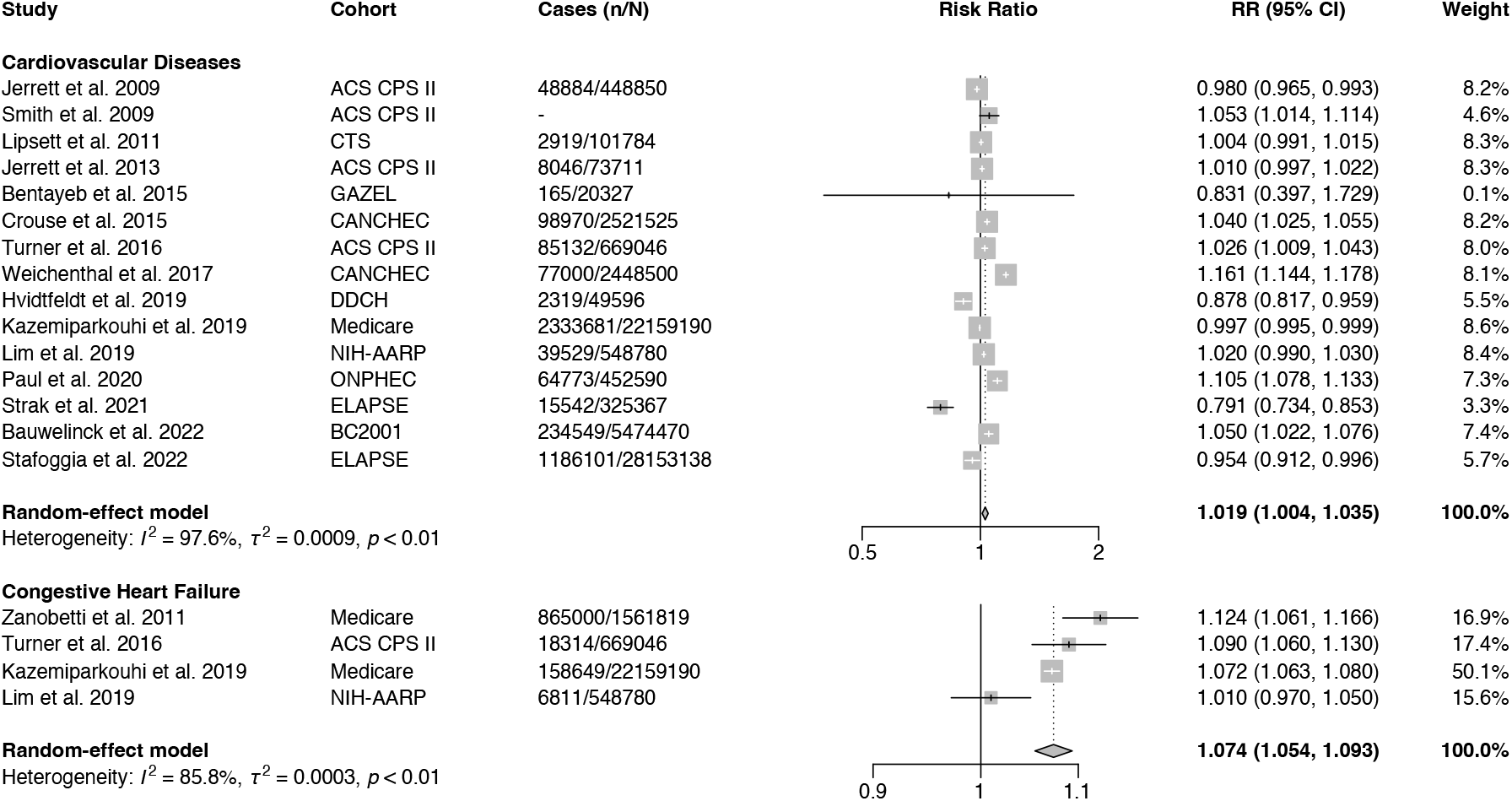
Pooled estimates of cardiovascular diseases and congestive heart failure mortality risk associated with every 10-ppbV incremental O_3_ exposure by 6mDMA8 metric.

#### 3.3.4 Other mortality causes

The other cause-specific mortality risks attributable to long-term O_3_ exposure were not statistically significant (**Figure 6**), as IHD mortality risk pooled from 10 studies was RR = 1.012 (95% CI: 0.987–1.039, I^2^ = 98.7%), CEVD mortality risk pooled from 6 studies was RR = 0.993 (95% CI: 0.979–1.008, I^2^ = 80.6%), and LC mortality risk pooled from 13 studies was RR = 0.966 (95% CI: 0.926–1.007, I^2^ = 84.2%). For all 8 studied mortality causes, we also provided pooled risks by 3 more widely used metrics (i.e. 6mDA24, ADMA8, and ADA24) besides 6mDMA8, as listed in **Table 3** for reference.

**Table 3.** Pooled RRs for long-term 10-ppbV incremental O_3_-exposure attributable multi-cause mortalities by 4 most widely used metrics and crude risks without metric harmonisation.

| Mortality causes | 6mDMA8 | 6mDA24 | ADMA8 | ADA24 | Crude |
| --- | --- | --- | --- | --- | --- |
| All causes (*n* = 23) | 1.014 (1.009, 1.019) | 1.023 (1.014, 1.032) | 1.016 (1.010, 1.022) | 1.027 (1.017, 1.037) | 1.017 (1.011, 1.023) |
| Respiratory diseases (*n* = 16) | 1.025 (1.010, 1.040) | 1.042 (1.016, 1.069) | 1.029 (1.011, 1.047) | 1.049 (1.019, 1.081) | 1.031 (1.017, 1.046) |
| Chronic obstructive pulmonary disease (*n* = 7) | 1.056 (1.029, 1.084) | 1.098 (1.050, 1.149) | 1.066 (1.034, 1.098) | 1.116 (1.058, 1.176) | 1.055 (1.032, 1.078) |
| Cardiovascular diseases (*n* = 15) | 1.019 (1.004, 1.035) | 1.033 (1.006, 1.061) | 1.022 (1.004, 1.041) | 1.038 (1.007, 1.071) | 1.024 (1.009, 1.038) |
| Ischaemic heart disease (*n* = 10) | 1.012 (0.987, 1.039) | 1.021 (0.977, 1.067) | 1.014 (0.984, 1.045) | 1.024 (0.973, 1.078) | 1.017 (0.994, 1.041) |
| Congestive heart failure (*n* = 4) | 1.074 (1.054, 1.093) | 1.130 (1.094, 1.168) | 1.086 (1.063, 1.110) | 1.155 (1.110, 1.198) | 1.083 (1.059, 1.107) |
| Cerebrovascular diseases (*n* = 6) | 0.993 (0.979, 1.008) | 0.988 (0.964, 1.013) | 0.992 (0.976, 1.009) | 0.986 (0.958, 1.015) | 0.992 (0.979, 1.006) |
| Lung cancer (*n* = 12) | 0.966 (0.926, 1.007) | 0.943 (0.878, 1.012) | 0.960 (0.915, 1.008) | 0.933 (0.859, 1.014) | 0.960 (0.909, 1.013) |

**Figure 6.**
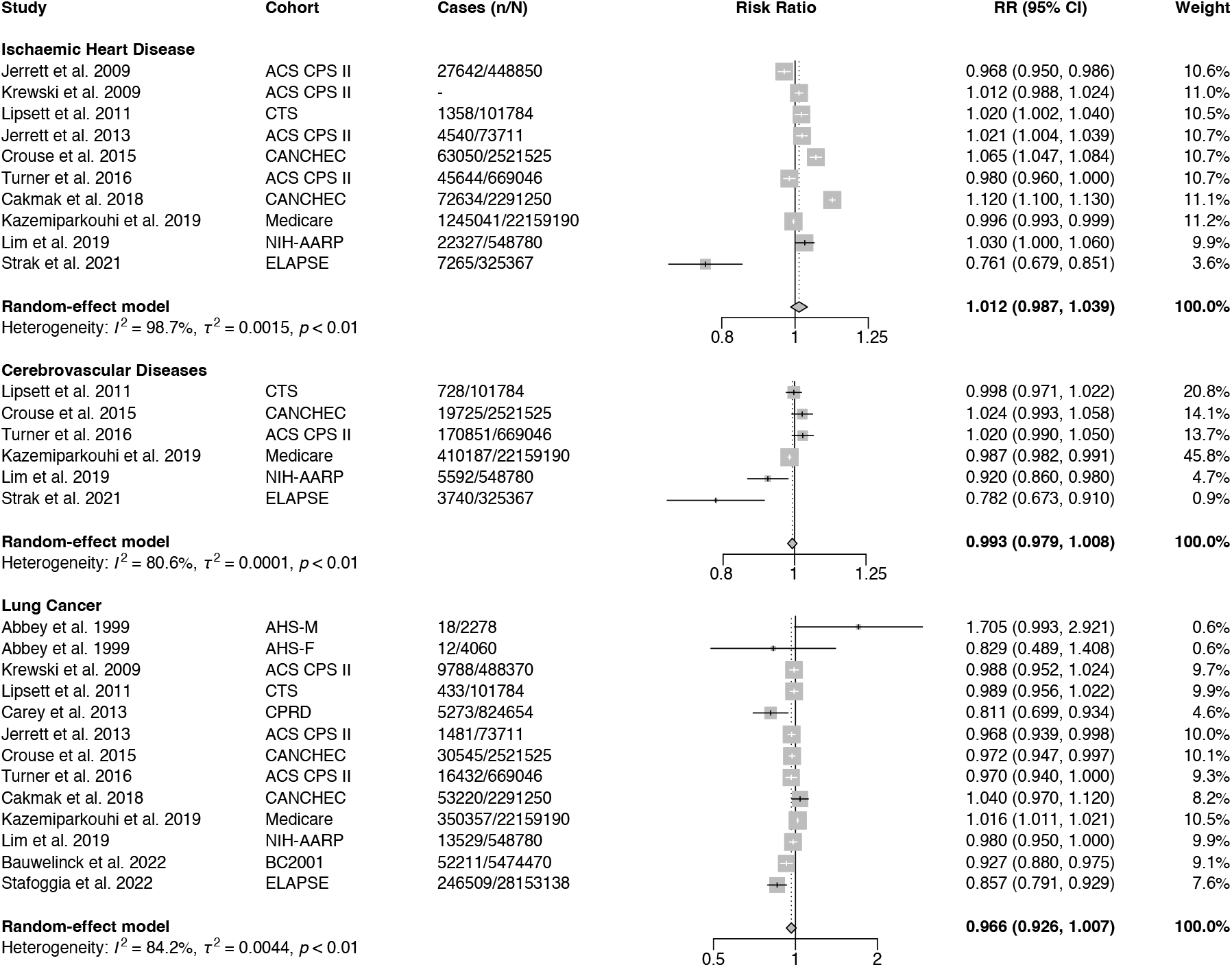
Pooled estimates of ischaemic heart disease, cerebrovascular diseases, lung cancer, ischaemic stroke, and pneumonia mortality risks associated with every 10-ppbV incremental O_3_ exposure by 6mDMA8 metric.

### 3.4 Study assessment

All 25 studies included into our final meta-analysis were rated to be above “Fair” (14 as “Fair” and 11 as “Good”) by the Quality Assessment Tool for Observational Cohort Studies, as listed in Table S4. All studies well met 10 out of 14 assessment items, while 9 studies did not sufficiently clarify the participant exclusion criteria; 2 re-analysis study reports did not clearly state the O_3_ exposures;^41, 42^ 2 studies were of relatively insufficient follow-up durations (e.g. less than 5 years) to observe the outcomes resulting from long-term exposure;^45, 63^ and 10 studies were of methodological deficiencies in individual exposure assignment,^38-46, 55^ most of which were conducted before 2013 when data assimilation techniques were not maturely developed to fuse observations with other full spatial coverage products such as satellite-based remote sensing and atmospheric mechanistic simulations. The satisfactory assessment results overall indicated inconspicuous risks of bias, laying the reliable foundation for meta-analyses.

Tables S5 displayed GRADE epidemiological evidence assessment results for each mortality cause from all involved corresponding studies. In brief, the overall judgements for all-cause, respiratory, cardiovascular, ischaemic heart disease, congestive heart failure, and lung cancer mortality risks were “High”, while the rating for the rest 2 cause-specific mortality risks (COPD and cerebrovascular diseases) were both “Moderate”.

Inconsistence of the risk directions (i.e. positive or negative associations) was the most common reason for downgrading, except for the CHF-induced mortality. There were 6 studies having reported the O_3_-mortality exposure-response trends to support the additional risks, as an assessment upgrading item for the pooled RRs of all-cause, respiratory and cardiovascular mortality. Cakmak et al. (2018) spotted higher RRs after adjusting the confounder compared to the crude values,^53^ which gave prominence to the positive risk associations and thus correspondingly upgraded the rating for all-cause, ischaemic heart disease, and lung cancer mortalities. No substantial positive publication biases were found based on the collected evidences.

### 3.5 Sensitivity analysis

Leave-one-out sensitivity analyses showed stable risk estimates as summarised in Table S6, except for the lung cancer mortality risks after eliminating Kazemiparkouhi et al. (2019), the only study reporting positive risk association,^55^ while the rest 11 studies concluded insignificant risks or even protective effects. Since the metric harmonisation in our study was an innovative attempt, we provided both metric-adjusted and unadjusted crude results for reference as presented in **Table 3**. The crude results were pooled from the originally reported relative risk values only unified into per 10-ppbV incremental exposure, without being transformed into any metrics for congruity. Along with the meta-analyses on all qualified studies, the relative risks were also pooled by keeping only one latest study with the largest population for each separate cohort, as summarised in Table S7. Under this circumstance, the pooled unit incremental mortality risks with every 10-ppbV incremental O_3_ exposure by 6mMDA8 metric were RR = 1.008 (95% CI: 1.006– 1.009, I^2^ = 82.6%) for all causes, RR = 1.034 (95% CI: 1.017–1.050, I^2^ = 81.7%) for all respiratory diseases, RR = 1.060 (95% CI: 1.040–1.080, I^2^ = 90.2%) for COPD, RR = 1.032 (95% CI: 1.010–1.055, I^2^ = 98.2%) for all cardiovascular diseases, RR = 1.008 (95% CI: 0.973–1.045, I^2^ = 99.2%) for ischaemic heart disease, and RR = 0.966 (95% CI: 0.931–1.002, I^2^ = 83.8%) for lung cancer. Studies for mortality risks of cerebrovascular diseases and congestive heart failure were respectively conducted on different cohorts, and hence such supplementary analysis was unnecessary.

## 4. DISCUSSION

### 4.1 Improvements as an updated review

This work improves on 2 previous high-quality reviews^15, 16^ by covering up-to-date peer-reviewed studies, and expanding the O_3_-exposure associated causes of mortality into wider range of categories. It is the first systematic review of the association between long-term O_3_ exposure and cause-specific mortality highlighting the issue of inconsistent use of exposure metrics to our best knowledge. Since tropospheric O_3_ is a photochemical pollutant which largely depends on solar radiation, the surface O_3_ concentrations can vary drastically between day and night, as well as warmer and cooler seasons. We pointed out that a 10-ppbV increase in annual daily 24-hour average concentration (ADA24) is more constrained in magnitude than a 10-ppbV increase in warm-season daily 8-hour maximum average concentration (6mDMA8) owing to the wider variability in the range of the latter metric. Taking the observations by TOAR and CNEMC *in situ* monitoring networks during 1990-2019 as an example, the surface O_3_ concentrations were 27.6 ± 6.1 (IQR: 24.1–31.0) ppbV by ADA24, while correspondingly 53.1 ± 10.6 (IQR: 47.7–61.4) ppbV by 6mDMA8, which indicated a 10-ppbV change fell below the IQR by the 6mDMA8, but could exceed the IQR using the ADA24 metric. This was why we believe adjusting the exposure metrics was necessary for O_3_ exposure-attributable health risk meta-analysis.

We also put forward a feasible approach to mutually convert the O_3_ exposure concentrations and corresponding risk strengths in various metrics by non-intercept linear projections following the operational suggestions from EPA,^32^ but update the linear conversion factors based on global *in situ* surface O_3_ observations during 1990-2019. The methodological innovation took advantage of multi-dimensional information from the original studies, which could inspire further observation collections and researches for corroborations and improvements.

### 4.2 Metric relevant issues

Although linear coefficients were applied onto the cross-metric conversions, irreducible noises still existed given the high root mean squared errors (RMSE) as shown in **Figure 2**, which exposed the limitation of risk strength adjustment into the same exposure metric by simple linear conversion, as the actual cross-metric relationships could be way more complicated. However, there was no other way but using the linear conversion coefficients as surrogates to unify the RRs by different metric reported in original studies, and thus to avoid uncertainties brought by the conversion of metrics, using a promissory consistent exposure metric or estimating the unit excess RRs in multiple metrics would be highly advocated in future long-term O_3_-exposure epidemiology studies.

Such linear conversion of risk associations could be validated by Kazemiparkouhi et al. (2020),^55^ where multiple metrics were applied to estimate the mortality risks. For COPD mortality, the RR was 1.072 (95% CI: 1.067–1.077) by 6mDMA1 for every 10-ppbV additional exposure; and after converting into 6mDMA8 metric using the linear coefficient 0.831 (**Figure 2**), the estimated RR was 1.087 (95% CI: 1.081–1.093), close to the literature reported 1.084 (95% CI: 1.079–1.089),^55^ which justified our linear conversion method. Converting Cross-metric linear conversions would not change the risk association direction, but using different exposure metrics when estimating the O_3_-exposure attributable mortality risks could potentially cause discrepancies. For an instance, Kazemiparkouhi et al. (2020) concluded excess hazards of long-term O_3_ exposure on all-cause mortality using 6mDMA1 and 6mDMA8 as quantitative metrics, but 6mDA24 led to a specious prevention effect (RR = 0.990, 95% CI: 0.988–0.991), which should be attributed to the existence of a theoretical exposure safety level for O_3_ below which no negative health effects should occur. Under this circumstance, lower-level metrics (e.g. ADA24) by averaging the peak O_3_ exposures might obscure the effective doses above the threshold, and also reduce the signal-to-noise ratios, so that were of lower credibility in recognising hazardous population exposures than higher-level metrics (e.g. 6mDMA8).

Data mining techniques were able to realise high-accuracy predictions of surface O_3_ concentrations, but errors were never avoidable. Carey et al. (2013) used a basic IDW spatial interpolation approach to obtain the surface O_3_ concentrations where the R^2^ were 0.24–0.76,^45^ while years later Di et al. (2017) applied an ensemble learning approach, achieving R^2^ = 0.80, RMSE = 2.91 ppbV.^51^ Carey et al. (2013) reported the IQR of O_3_ exposure concentrations as 3.0 ppbV, which was comparable to the RMSE of Di et al. (2017).^51^ Besides, lower R^2^ could be accompanied with higher prediction errors, which might have concealed the highest and lowest quartiles, and led to failures in distinguishing the population-level exposures. This concern had been reflected in our subgroup meta-analysis by exposure metrics, that lower-level metrics were more inclined to report insignificant risks, which also cast sceptics on the reliability of studies covering narrow exposure variabilities. We therefore are in favour of the Lancet suggestions to use peak metrics to quantify the long-term O_3_ exposure such as 6mDMA8, and also speak highly of the state-of-the-art data techniques to reduce errors in O_3_ concentration prediction, so as to make a distinction between the high- and low-exposure populations.

### 4.3 Pathogenesis supports

Atkinson et al. (2016) concluded insignificant pooled risks for long-term O_3_ exposure associated all-cause and respiratory mortality,^16^ which contradicted our results. It should mainly be ascribed to the heterogeneity between the more recent studies and earlier ones. The majority of studies collected in Atkinson et al. (2016) applied primitive statistical methods (i.e. nearest neighbourhood matching, IDW and ordinary kriging interpolation) for individual exposure assignment, which might have weakened the individual-level exposure distinguishment. In addition, some studies using ADA24 as the exposure metric could have also obscured the significance of associations.^43, 45, 46^ In contrast, studies after 2016 more frequently applied advanced numerical simulation models and data assimilation techniques to increase the precision of population exposure assessment; and most of them used 6mDMA8 metric to foreground the high exposures.^50, 52, 53, 55-57^ These recent studies stuck out the significant O_3_-mortality associations.

From another aspect, pathological mechanisms had been at least partially ascertained by laboratorial experiments. The inhaled O_3_ could constrict the muscles in the airways leading to shortness of breath, and damage the lining with inflammation.^64^ Long-term O_3_ exposure could increase the oxidative stress in the cardiovascular system,^65^ and cause progressive thickening of the carotid arteries to restrict cerebral blood supply.^66^ Additionally, strong associations had already been found between short-term O_3_ exposure and a variety of cardiopulmonary symptoms as reported by a number of observational epidemiological studies,^6^ which also supported the long-term exposure effects, as it was unreasonable to presume no incremental risks by long-term exposure given the verified significant short-term effects. We thus were inclined to approve of the opinion that long-term O_3_ exposure would increase mortality risks in agreement with GBD report.^20^

To alleviate the population health loss resulting from O_3_ exposure, the U.S. EPA appealed for optimisations in real-time accessibility of air quality index, with which residents could be able to avoid unnecessary high pollution exposure (https://www.epa.gov/ground-level-ozone-pollution/health-effects-ozone-pollution). Appropriate diets and supplements including carotenoids, vitamin D and vitamin E were recognised to be preventive against air pollution induced respiratory damages, which was a practical protective measure for the vulnerable.^67^

### 4.4 Concentration-response relationship

Few studies had examined the concentration-response curves between long-term O_3_ exposure and mortality, and thus the threshold exposure level (also known as theoretical minimum risk exposure level, TMREL) below which no adverse health effects would be assumed to occur was still controversial. For all-cause mortality, Di et al. (2017) reported a safe exposure level as 30 ppbV by 6mDA24 metric (approximately as 49.9 ppbV by 6mDMA8),^51^ while Shi et al. (2022) suggested a lower level as 40 ppbV by 6mDMA8, both estimated from the Medicare beneficiary cohort.^58^ For respiratory mortality, Jerrett et al. (2009) tested the concentration-response relationships and estimated the threshold level as 60 ppbV by 6mDMA1 (49.9 ppbV by 6mDMA8),^40^ while Lim et al. (2019) failed to identify a significant threshold level.^56^ For cardiovascular mortality, Lim et al. (2019) showed no apparent health hazards below 45 ppbV by 6mDMA8,^56^ and Paul et al. (2020) prescribed a threshold level around 35 ppbV by 6mDMA8 metric for diabetic patients.^57^ These evidence-based threshold exposure levels were all no higher than the current standards, as 70 ppbV for daily maximum 8-hour exposure under NAAQS (The National Ambient Air Quality Standards regulated by the U.S. EPA)^68^ and 50 ppbV by warm-season DMA8 under WHO global air quality guidelines.^69^ However, whether the standard guidelines should be revised to be more strictly would require more further studies.

To synthesise epidemiological evidences, Burnett et al. (2014) developed an integrated exposure-response (IER) function-based curve-fitting method to pool the risk associations from multiple studies.^37^ We attempted to construct the IER for long-term O_3_ exposure associated mortalities in this review, with statistically reproduced exposure levels to enhance the curve fitting, as illustrated in Supplementary Texts S1-S3. The exposure imputing had revealed high reliability, but the high uncertainties of the IER curves could still not be addressed, which should be attributed to the limited effective epidemiological evidences. Empirically, this approach would require sufficient studies to cover a wide range of exposure levels, which had been frequently adopted for particulate matter exposure researches,^70-72^ but seldomly used for O_3_-health studies.^20^ The main probable reason might be that the population long-term O_3_ exposure levels would not be as comparably distinguishable to the particulate matters. In addition, a reasonable prescribed TMREL would be necessary to establish the IER curves,^37^ and hence the indeterminacy of the threshold level could exacerbate the uncertainties in the estimated concentration-response relationships. Therefore, more relevant studies on long-term O_3_ exposure associated risks are urgently appealed for, based on which discussions, optimisations, or corrections on our enhanced exposure-response curve-fitting methodologies, are encouraged.

### 4.5 Hierarchical classification of diseases

The causes of mortalities analysed in our study followed hierarchical subordinate relationships, as the all-cause mortality consisted of cardiovascular diseases, respiratory diseases, cancer and other causes; chronic obstructive pulmonary disease belonged to respiratory category; and ischaemic heart disease, stroke, congestive heart failure and other cerebrovascular diseases all subordinated to cardiovascular symptoms. On this occasion, estimating all O_3_-exposure induced mortalities could follow a bottom-up scheme by adding up subgroups of diseases. However, for the historical O_3_-associated mortalities, GBD attributed all O_3_-associated mortalities onto COPD-induced premature deaths,^20^ which we thought were of spaces for further explorations. Long-term O_3_ exposure had shown significant association with excess cardiovascular mortalities, and thus we should update the mortality estimations in further studies by including CVD altogether into consideration.

### 4.6 Application in mortality estimations

The widest applications of the estimated risk association strengths were to project how many people would be affected by long-term ambient O_3_ exposure. For example, Malley et al. (2017) estimated 1.23 (95% UI: 0.85–1.62) million respiratory deaths attributable to O_3_ exposure in 2010,^73^ using the risk strength by Turner et al. (2016) as HR = 1.12 (95% UI: 1.08–1.16).^50^ This estimation was much higher than the 2019 GBD report: 0.31 (95% UI: 0.15–0.49) million, as had been highlighted in another recent study,^25^ which should be attributed to the use of high HR value among all included studies. We had also found some other studies using one singular HR value for population risk estimations,^17, 74-77^ but we would still encourage further relevant studies to consider multi-study pooled RRs, which could effectively reduce the potential biases from a single study. The adaptability of the pooled RRs could be verified from the coverage of exposure levels, as the 25 studies identified in our review had embraced a wide range of exposure concentrations (Supplementary Text S1) to encompass the global surface O_3_ variability.^25^ On the other hand, the leave-one-out sensitivity analyses (Table S6) had revealed the robustness of the meta-analysis results when including sufficient numbers of studies, which was a circumstantial reflection for the representativeness of the synthesised risk association strengths. The annual GBD reports were also based on the generalisability presumption of the synthesised epidemiological evidences, but cohort-based researches in the unstudied regions are always appealed for to provide more convincing discoveries.

### 4.7 Limitations

Although the total number of studied participants for risk pooling was adequately high to ensure the statistical power, the cohort-based O_3_-health studies were factually rare according to our literature search, and thus long-term follow-up studies are urgently encouraged. Additionally, current literatures seldomly reported grouped RRs, which made meta-analyses by sub-categories (like gender, age, socio-economic status, smoking and alcohol history, etc.) unfeasible. Scarcity of credible evidences also restricted the effects of conventional approaches to construct exposure-response curves, and our methodological innovation would require further relevant studies for substantiation. The cross-metric linear conversion factors were estimated relying on observations from available sites, which however might not be sufficiently representative of the global residential areas, as observational sites in India, Africa, and Latin America were still sparse. With ever-increasing popularisation of the *in situ* monitoring networks, the cross-metric conversion factors might need calibration with more comprehensive observations, so that the pooled RRs should also be updated accordingly.

### 4.8 Further study suggestions

We suggest that further environmental epidemiology studies, especially long-term O_3_ exposure related researches, clearly report i) the methodologies to obtain ambient O_3_ concentrations, the spatiotemporal resolution, and prediction accuracy of the database; ii) the exposure metrics used for risk estimation; and iii) the statistical distribution of the O_3_ exposure concentrations. The data-oriented methodologies to accomplish full spatial coverage ambient air O_3_ concentrations for individual-level exposure assignment should be transparent as the construction credibility of air pollution concentration database should also be rigorously assessed, which were the foundation of epidemiological follow-up studies. We would advocate the report of exposure metrics in future O_3_-health studies so as to avoid confusions when comparing the risks with literature and conducting meta-regression; and according to the recent consensus, warm-season average (6mDMA8) shall be preferred as epidemiological study metrics.^19^ We recommend future studies estimate risks with multiple O_3_ metrics for reference; and describing the statistical distribution of the O_3_ exposure levels is another suggested element to assess the reliability of risk estimation models, which can also be useful in exposure-response tendency exploration. We also propose future cohort studies estimate subgroup-specific RRs which can be conducive to identify the vulnerable populations.

Our review highlights a deficiency existing in current environmental health research literatures, that studies on long-term O_3_ exposure health effects are still rather rare compared to particulate matter-based studies.^78^ Also, the meta-analysis results might be geographically-biased, since large-scale O_3_ exposure health risk studies till 2022 did not cover Asia, Africa or Latin America regions. However, there are some thriving cohorts such as the Multi-Country Multi-City (MCC) Collaborative Research Network covering over 22 countries or regions,^79^ and the China Kadoorie Biobank (CKB) enrolling over 0.5 million people,^80^ working on environmental exposure projects. We are optimistic that more research will come out to fill in the literature gap of multi-region population-based studies.

## 5. CONCLUSION

Our state-of-the-science systematic review has summarised cohort studies exploring the associations between long-term ambient O_3_ exposure and multi-cause mortality risks. Current studies support O_3_-exposure attributable additional mortalities caused from all causes, respiratory diseases, chronic obstructive pulmonary disease, cardiovascular diseases, and congestive heart failure, but no significant mortality risks are found for ischaemic heart diseases, all-type cerebrovascular diseases, and lung cancer. Exposure metrics are crucial for health risk estimations of long-term O_3_ exposure and meta-analysis to pool the multi-study risks, for which we develop a linear conversion approach to harmonise the different metrics. Further researches on long-term O_3_ observations and exposure-induced mortalities are encouraged to corroborate or contradict our linear conversion factors and meta-analysis results by providing more solid evidences, so as to strengthen the O_3_-health literatures.

## Data Availability

The surface O_3_ observations are archived in the Tropospheric Ozone Assessment Report (TOAR, https://b2share.fz-juelich.de/communities/TOAR) repository and China National Environmental Monitoring Centre (CNEMC, http://www.cnemc.cn/en/) repository, which are accessible to the public. The cohort-based long-term O_3_ exposure-associated cause-specific mortality risks are all available in the main text or supplementary materials of the selected studies. 

## Competing Interest Statement

All authors declare: no support from any organisation for the submitted work; no financial relationships with any organisations that might have an interest in the submitted work in the previous three years; and no other relationships or activities that could appear to have influenced the submitted work.

## Author Contributions

ATA and YG conceived the idea for the review; HZS, CL and PY performed the literature search; HZS and PY conducted statistical analyses; HZS, ATA, YG, PY, SH, JM, HS and LY contributed to discussions; HZS wrote the article; and MW examined the languages. HZS is the guarantor who accepted the full responsibility for the finished article, owned full access to all relevant data, and controlled the decision to publish. The two joint corresponding authors, ATA and YG, attests that all listed authors meet authorship criteria and that no others meeting the criteria have been omitted. The guarantor, HZS, affirms that the manuscript is an honest, accurate, and transparent account of the study being reported, and no important aspects of the study have been omitted. No ethical approval is needed for a systematic review and meta-analysis.

## Acknowledgement

This study is funded by UK Natural Environment Research Council (NERC), UK National Centre for Atmospheric Science (NCAS), Australian Research Council (DP210102076) and Australian National Health and Medical Research Council (APP2000581). HZS, MW, and SH receive funding from Engineering and Physical Sciences Research Council (EPSRC) via the UK Research and Innovation (UKRI) Centre for Doctoral Training in Application of Artificial Intelligence to the study of Environmental Risks (AI4ER, EP/S022961/1). ATA acknowledges funding from NERC (NE/P016383/1) and through the Met Office UKRI Clean Air Programme. YG is supported by a Career Development Fellowship of the Australian National Health and Medical Research Council (APP1163693). All contents in this study are solely the responsibility of the grantees and do not represent the official views of the supporting agencies.

Special appreciations to Dr Xiao Lu (School of Atmospheric Sciences, Sun Yat-sen University) for his insightful discussion on the quality control of TOAR products, Dr Liuhua Shi (Rollins School of Public Health, Emory University) for her supplementary information on Medicare beneficiary cohort information, and 4 anonymous reviewers together with the editor for their meticulous efforts in improving the manuscript.

## Supplementary Information

Further detailed information can be found in the **Supplementary Materials** (PDF) consisting of 25 pages with 5 sections of texts as “imputation procedures for exposure distribution” (S1), “enhanced integrated exposure-response curve-fitting” (S2), “demonstrative procedures of enhanced exposure-response trend curve-fitting” (S3), “undefined metric imputation” (S4), and “interpretation and procedure of cross-metric linear conversion” (S5), together with 6 tables and 7 figures to strengthen the results and discussions presented in the main text. A **PRISMA Checklist** (PDF) was provided to verify the integrity of this study.

## Notes

### Competing Interest Statement

The authors have declared no competing interest.

## REFERENCES

1. Fu, B.; Gasser, T.; Li, B.; Tao, S.; Ciais, P.; Piao, S.; Balkanski, Y.; Li, W.; Yin, T.; Han, L., Short-lived climate forcers have long-term climate impacts via the carbon-climate feedback. Nat Clim Change 2020, 10, (9), 851–855.

2. Wilson, S. R.; Madronich, S.; Longstreth, J. D.; Solomon, K. R., Interactive effects of changing stratospheric ozone and climate on tropospheric composition and air quality, and the consequences for human and ecosystem health. Photochem Photobiol Sci 2019, 18, (3), 775–803.

3. Stolarski, R. S., The Antarctic ozone hole. Sci Am 1988, 258, (1), 30–37.

4. Ghude, S. D.; Jena, C.; Chate, D. M.; Beig, G.; Pfister, G. G.; Kumar, R.; Ramanathan, V., Reductions in India’s crop yield due to ozone. Geophys Res Lett 2014, 41, (15), 5685–5691.

5. Zhang, F.; Zhang, H.; Wu, C.; Zhang, M.; Feng, H.; Li, D.; Zhu, W., Acute effects of ambient air pollution on clinic visits of college students for upper respiratory tract infection in Wuhan, China. Environ Sci Pollut Res Int 2021, 1–11.

6. Zheng, X.-Y.; Orellano, P.; Lin, H.-L.; Jiang, M.; Guan, W.-J., Short-term exposure to ozone, nitrogen dioxide, and sulphur dioxide and emergency department visits and hospital admissions due to asthma: A systematic review and meta-analysis. Environ Int 2021, 150, 106435.

7. Liu, Y.; Pan, J.; Fan, C.; Xu, R.; Wang, Y.; Xu, C.; Xie, S.; Zhang, H.; Cui, X.; Peng, Z.; Shi, C.; Zhang, Y.; Sun, H.; Zhou, Y.; Zhang, L., Short-Term Exposure to Ambient Air Pollution and Mortality From Myocardial Infarction. J Am Coll Cardiol 2021, 77, (3), 271–281.

8. Zhao, R.; Chen, S.; Wang, W.; Huang, J.; Wang, K.; Liu, L.; Wei, S., The impact of short-term exposure to air pollutants on the onset of out-of-hospital cardiac arrest: A systematic review and meta-analysis. Int J Cardiol 2017, 226, 110–117.

9. Han, C.; Lu, Y.; Cheng, H.; Wang, C.; Chan, P., The impact of long-term exposure to ambient air pollution and second-hand smoke on the onset of Parkinson disease: a review and meta-analysis. Public Health 2020, 179, 100–110.

10. Li, J.; Huang, J.; Cao, R.; Yin, P.; Wang, L.; Liu, Y.; Pan, X.; Li, G.; Zhou, M., The association between ozone and years of life lost from stroke, 2013-2017: A retrospective regression analysis in 48 major Chinese cities. J Hazard Mater 2021, 405, 124220.

11. Liu, G.; Sun, B.; Yu, L.; Chen, J.; Han, B.; Li, Y.; Chen, J., The Gender-Based Differences in Vulnerability to Ambient Air Pollution and Cerebrovascular Disease Mortality: Evidences Based on 26781 Deaths. Glob Heart 2020, 15, (1), 46.

12. Sun, Z.; Yang, L.; Bai, X.; Du, W.; Shen, G.; Fei, J.; Wang, Y.; Chen, A.; Chen, Y.; Zhao, M., Maternal ambient air pollution exposure with spatial-temporal variations and preterm birth risk assessment during 2013-2017 in Zhejiang Province, China. Environ Int 2019, 133, (Pt B), 105242.

13. Global Burden of Disease Collaborative Network, Global Burden of Disease Study 2019 (GBD 2019) Results. In Institute for Health Metrics and Evaluation (IHME): Seattle, United States, 2020.

14. Niu, Z.; Liu, F.; Yu, H.; Wu, S.; Xiang, H., Association between exposure to ambient air pollution and hospital admission, incidence, and mortality of stroke: an updated systematic review and meta-analysis of more than 23 million participants. Environ Health Prev Med 2021, 26, (1), 1–14.

15. Huangfu, P.; Atkinson, R., Long-term exposure to NO_2_ and O_3_ and all-cause and respiratory mortality: A systematic review and meta-analysis. Environ Int 2020, 144, 105998.

16. Atkinson, R. W.; Butland, B. K.; Dimitroulopoulou, C.; Heal, M. R.; Stedman, J. R.; Carslaw, N.; Jarvis, D.; Heaviside, C.; Vardoulakis, S.; Walton, H.; Anderson, H. R., Long-term exposure to ambient ozone and mortality: a quantitative systematic review and meta-analysis of evidence from cohort studies. BMJ Open 2016, 6, (2), e009493.

17. Shen, H.; Sun, Z.; Chen, Y.; Russell, A. G.; Hu, Y.; Odman, M. T.; Qian, Y.; Archibald, A. T.; Tao, S., Novel Method for Ozone Isopleth Construction and Diagnosis for the Ozone Control Strategy of Chinese Cities. Environ Sci Technol 2021, 55, (23), 15625–15636.

18. Sun, Z.; Archibald, A. T., Multi-stage Ensemble-learning-based Model Fusion for Surface Ozone Simulations: A Focus on CMIP6 Models. Environmental Science and Ecotechnology 2021, 8, 100124.

19. Schultz, M. G.; Schroder, S.; Lyapina, O.; Cooper, O. R.; Galbally, I.; Petropavlovskikh, I.; von Schneidemesser, E.; Tanimoto, H.; Elshorbany, Y.; Naja, M.; Seguel, R. J.; Dauert, U.; Eckhardt, P.; Feigenspan, S.; Fiebig, M.; Hjellbrekke, A. G.; Hong, Y. D.; Kjeld, P. C.; Koide, H.; Lear, G.; Tarasick, D.; Ueno, M.; Wallasch, M.; Baumgardner, D.; Chuang, M. T.; Gillett, R.; Lee, M.; Molloy, S.; Moolla, R.; Wang, T.; Sharps, K.; Adame, J. A.; Ancellet, G.; Apadula, F.; Artaxo, P.; Barlasina, M. E.; Bogucka, M.; Bonasoni, P.; Chang, L.; Colomb, A.; Cuevas-Agullo, E.; Cupeiro, M.; Degorska, A.; Ding, A. J.; FrHlich, M.; Frolova, M.; Gadhavi, H.; Gheusi, F.; Gilge, S.; Gonzalez, M. Y.; Gros, V.; Hamad, S. H.; Helmig, D.; Henriques, D.; Hermansen, O.; Holla, R.; Hueber, J.; Im, U.; Jaffe, D. A.; Komala, N.; Kubistin, D.; Lam, K. S.; Laurila, T.; Lee, H.; Levy, I.; Mazzoleni, C.; Mazzoleni, L. R.; McClure-Begley, A.; Mohamad, M.; Murovec, M.; Navarro-Comas, M.; Nicodim, F.; Parrish, D.; Read, K. A.; Reid, N.; Ries, N. R. L.; Saxena, P.; Schwab, J. J.; Scorgie, Y.; Senik, I.; Simmonds, P.; Sinha, V.; Skorokhod, A. I.; Spain, G.; Spangl, W.; Spoor, R.; Springston, S. R.; Steer, K.; Steinbacher, M.; Suharguniyawan, E.; Torre, P.; Trickl, T.; Lin, W. L.; Weller, R.; Xu, X. B.; Xue, L. K.; Ma, Z. Q., Tropospheric Ozone Assessment Report: Database and metrics data of global surface ozone observations. Elem Sci Anth 2017, 5, 58–83.

20. Institute for Health Metrics and Evaluation (IHME), GBD 2019 Cause and Risk Summary: Ambient Ozone Pollution. In IHME, University of Washington: Seattle, USA, 2020.

21. Guyatt, G. H.; Oxman, A. D.; Vist, G. E.; Kunz, R.; Falck-Ytter, Y.; Alonso-Coello, P.; Schunemann, H. J.; Group, G. W., GRADE: an emerging consensus on rating quality of evidence and strength of recommendations. BMJ 2008, 336, (7650), 924–6.

22. Guyatt, G.; Oxman, A. D.; Akl, E. A.; Kunz, R.; Vist, G.; Brozek, J.; Norris, S.; Falck-Ytter, Y.; Glasziou, P.; DeBeer, H.; Jaeschke, R.; Rind, D.; Meerpohl, J.; Dahm, P.; Schunemann, H. J., GRADE guidelines: 1. Introduction-GRADE evidence profiles and summary of findings tables. J Clin Epidemiol 2011, 64, (4), 383–94.

23. Egger, M.; Davey Smith, G.; Schneider, M.; Minder, C., Bias in meta-analysis detected by a simple, graphical test. BMJ 1997, 315, (7109), 629–34.

24. Duval, S.; Tweedie, R., A nonparametric “trim and fill” method of accounting for publication bias in meta-analysis. J Am Stat Assoc 2000, 95, (449), 89–98.

25. Sun, H. Z.; Shin, Y. M.; Xia, M.; Ke, S.; Yuan, L.; Guo, Y.; Archibald, A. T., Spatial Resolved Surface Ozone with Urban and Rural Differentiation during 1990-2019: A Space-time Bayesian Neural Network Downscaler. Environ Sci Technol 2021.

26. Krupa, S.; McGrath, M. T.; Andersen, C. P.; Booker, F. L.; Burkey, K. O.; Chappelka, A. H.; Chevone, B. I.; Pell, E. J.; Zilinskas, B. A., Ambient Ozone and Plant Health. Plant Dis 2001, 85, (1), 4–12.

27. Finlayson-Pitts, B.; Pitts Jr, J., Atmospheric chemistry of tropospheric ozone formation: scientific and regulatory implications. Air & Waste 1993, 43, (8), 1091–1100.

28. Fleming, Z. L.; Doherty, R. M.; Von Schneidemesser, E.; Malley, C. S.; Cooper, O. R.; Pinto, J. P.; Colette, A.; Xu, X.; Simpson, D.; Schultz, M. G.; Lefohn, A. S.; Hamad, S.; Moolla, R.; Solberg, S.; Feng, Z., Tropospheric Ozone Assessment Report: Present-day ozone distribution and trends relevant to human health. Elem Sci Anth 2018, 6, 12.

29. Griffiths, P. T.; Murray, L. T.; Zeng, G.; Archibald, A. T.; Emmons, L. K.; Galbally, I.; Hassler, B.; Horowitz, L. W.; Keeble, J.; Liu, J.; Moeini, O.; Naik, V.; amp; apos; Connor, F. M.; Shin, Y. M.; Tarasick, D.; Tilmes, S.; Turnock, S. T.; Wild, O.; Young, P. J.; Zanis, P., Tropospheric ozone in CMIP6 Simulations. Atmos Chem Phys 2021, 21, (5), 4187–4218.

30. Archibald, A. T.; O’Connor, F. M.; Abraham, N. L.; Archer-Nicholls, S.; Chipperfield, M. P.; Dalvi, M.; Folberth, G. A.; Dennison, F.; Dhomse, S. S.; Griffiths, P. T.; Hardacre, C.; Hewitt, A. J.; Hill, R.; Johnson, C. E.; Keeble, J.; Köhler, M. O.; Morgenstern, O.; Mulchay, J. P.; Ordóñez, C.; Pope, R. J.; Rumbold, S.; Russo, M. R.; Savage, N.; Sellar, A.; Stringer, M.; Turnock, S.; Wild, O.; Zeng, G., Description and evaluation of the UKCA stratosphere-troposphere chemistry scheme (StratTrop vn 1.0) implemented in UKESM1. Geosci Model Dev 2019, 13, (3), 1223–1266.

31. Tong, L.; Zhang, H.; Yu, J.; He, M.; Xu, N.; Zhang, J.; Qian, F.; Feng, J.; Xiao, H., Characteristics of surface ozone and nitrogen oxides at urban, suburban and rural sites in Ningbo, China. Atmos Res 2017, 187, 57–68.

32. U.S. Epa Air Quality Criteria For Ozone and Related Photochemical Oxidants (Final Report, 2006); U.S. Environmental Protection Agency: Washington, DC, 2006.

33. Tarasick, D.; Galbally, I. E.; Cooper, O. R.; Schultz, M. G.; Ancellet, G.; Leblanc, T.; Wallington, T. J.; Ziemke, J.; Liu, X.; Steinbacher, M.; Staehelin, J.; Vigouroux, C.; Hannigan, J. W.; Garcia, O.; Foret, G.; Zanis, P.; Weatherhead, E.; Petropavlovskikh, I.; Worden, H.; Osman, M.; Liu, J.; Chang, K. L.; Gaudel, A.; Lin, M. Y.; Granados-Munoz, M.; Thompson, A. M.; Oltmans, S. J.; Cuesta, J.; Dufour, G.; Thouret, V.; Hassler, B.; Trickl, T.; Neu, J. L., Tropospheric Ozone Assessment Report: Tropospheric ozone from 1877 to 2016, observed levels, trends and uncertainties. Elem Sci Anth 2019, 7, 39.

34. Lu, X.; Hong, J. Y.; Zhang, L.; Cooper, O. R.; Schultz, M. G.; Xu, X. B.; Wang, T.; Gao, M.; Zhao, Y. H.; Zhang, Y. H., Severe Surface Ozone Pollution in China: A Global Perspective. Environ Sci Technol Lett 2018, 5, (8), 487–494.

35. Hunter, J. E.; Schmidt, F. L., Meta-analysis. In Advances in educational and psychological testing: Theory and applications, Springer: 1991; pp 157–183.

36. Higgins, J. P.; Thompson, S. G.; Deeks, J. J.; Altman, D. G., Measuring inconsistency in meta-analyses. BMJ 2003, 327, (7414), 557–60.

37. Burnett, R. T.; Pope, C. A., 3rd; Ezzati, M.; Olives, C.; Lim, S. S.; Mehta, S.; Shin, H. H.; Singh, G.; Hubbell, B.; Brauer, M.; Anderson, H. R.; Smith, K. R.; Balmes, J. R.; Bruce, N. G.; Kan, H.; Laden, F.; Pruss-Ustun, A.; Turner, M. C.; Gapstur, S. M.; Diver, W. R.; Cohen, A., An integrated risk function for estimating the global burden of disease attributable to ambient fine particulate matter exposure. Environ Health Perspect 2014, 122, (4), 397–403.

38. Abbey, D. E.; Nishino, N.; McDonnell, W. F.; Burchette, R. J.; Knutsen, S. F.; Lawrence Beeson, W.; Yang, J. X., Long-term inhalable particles and other air pollutants related to mortality in nonsmokers. Am J Respir Crit Care Med 1999, 159, (2), 373–82.

39. Lipfert, F. W.; Wyzga, R. E.; Baty, J. D.; Miller, J. P., Traffic density as a surrogate measure of environmental exposures in studies of air pollution health effects: Long-term mortality in a cohort of US veterans. Atmos Environ 2006, 40, (1), 154–169.

40. Jerrett, M.; Burnett, R. T.; Pope, C. A., 3rd; Ito, K.; Thurston, G.; Krewski, D.; Shi, Y.; Calle, E.; Thun, M., Long-term ozone exposure and mortality. N Engl J Med 2009, 360, (11), 1085–95.

41. Krewski, D.; Jerrett, M.; Burnett, R. T.; Ma, R.; Hughes, E.; Shi, Y.; Turner, M. C.; Pope III, C. A.; Thurston, G.; Calle, E. E., Extended follow-up and spatial analysis of the American Cancer Society study linking particulate air pollution and mortality. Health Effects Institute Boston, MA: 2009.

42. Smith, K. R.; Jerrett, M.; Anderson, H. R.; Burnett, R. T.; Stone, V.; Derwent, R.; Atkinson, R. W.; Cohen, A.; Shonkoff, S. B.; Krewski, D.; Pope, C. A., 3rd; Thun, M. J.; Thurston, G., Public health benefits of strategies to reduce greenhouse-gas emissions: health implications of short-lived greenhouse pollutants. Lancet 2009, 374, (9707), 2091–2103.

43. Lipsett, M. J.; Ostro, B. D.; Reynolds, P.; Goldberg, D.; Hertz, A.; Jerrett, M.; Smith, D. F.; Garcia, C.; Chang, E. T.; Bernstein, L., Long-term exposure to air pollution and cardiorespiratory disease in the California teachers study cohort. Am J Respir Crit Care Med 2011, 184, (7), 828–35.

44. Zanobetti, A.; Schwartz, J., Ozone and survival in four cohorts with potentially predisposing diseases. Am J Respir Crit Care Med 2011, 184, (7), 836–41.

45. Carey, I. M.; Atkinson, R. W.; Kent, A. J.; van Staa, T.; Cook, D. G.; Anderson, H. R., Mortality associations with long-term exposure to outdoor air pollution in a national English cohort. Am J Respir Crit Care Med 2013, 187, (11), 1226–33.

46. Jerrett, M.; Burnett, R. T.; Beckerman, B. S.; Turner, M. C.; Krewski, D.; Thurston, G.; Martin, R. V.; van Donkelaar, A.; Hughes, E.; Shi, Y.; Gapstur, S. M.; Thun, M. J.; Pope, C. A., 3rd, Spatial analysis of air pollution and mortality in California. Am J Respir Crit Care Med 2013, 188, (5), 593–9.

47. Bentayeb, M.; Wagner, V.; Stempfelet, M.; Zins, M.; Goldberg, M.; Pascal, M.; Larrieu, S.; Beaudeau, P.; Cassadou, S.; Eilstein, D., Association between long-term exposure to air pollution and mortality in France: a 25-year follow-up study. Environ Int 2015, 85, 5–14.

48. Crouse, D. L.; Peters, P. A.; Hystad, P.; Brook, J. R.; van Donkelaar, A.; Martin, R. V.; Villeneuve, P. J.; Jerrett, M.; Goldberg, M. S.; Pope III, C. A., Ambient PM<sub>2.5</sub>, O<sub>3</sub>, and NO<sub>2 </sub>exposures and associations with mortality over 16 years of follow-up in the Canadian Census Health and Environment Cohort (CanCHEC). Environ Health Perspect 2015, 123, (11), 1180–1186.

49. Tonne, C.; Halonen, J. I.; Beevers, S. D.; Dajnak, D.; Gulliver, J.; Kelly, F. J.; Wilkinson, P.; Anderson, H. R., Long-term traffic air and noise pollution in relation to mortality and hospital readmission among myocardial infarction survivors. Int J Hyg Environ Health 2016, 219, (1), 72–78.

50. Turner, M. C.; Jerrett, M.; Pope, C. A., 3rd; Krewski, D.; Gapstur, S. M.; Diver, W. R.; Beckerman, B. S.; Marshall, J. D.; Su, J.; Crouse, D. L.; Burnett, R. T., Long-Term Ozone Exposure and Mortality in a Large Prospective Study. Am J Respir Crit Care Med 2016, 193, (10), 1134–42.

51. Di, Q.; Wang, Y.; Zanobetti, A.; Wang, Y.; Koutrakis, P.; Choirat, C.; Dominici, F.; Schwartz, J. D., Air Pollution and Mortality in the Medicare Population. N Engl J Med 2017, 376, (26), 2513–2522.

52. Weichenthal, S.; Pinault, L. L.; Burnett, R. T., Impact of oxidant gases on the relationship between outdoor fine particulate air pollution and nonaccidental, cardiovascular, and respiratory mortality. Sci Rep 2017, 7, (1), 1–10.

53. Cakmak, S.; Hebbern, C.; Pinault, L.; Lavigne, E.; Vanos, J.; Crouse, D. L.; Tjepkema, M., Associations between long-term PM<sub>2.5 </sub>and ozone exposure and mortality in the Canadian Census Health and Environment Cohort (CANCHEC), by spatial synoptic classification zone. Environ Int 2018, 111, 200–211.

54. Hvidtfeldt, U. A.; Sorensen, M.; Geels, C.; Ketzel, M.; Khan, J.; Tjonneland, A.; Overvad, K.; Brandt, J.; Raaschou-Nielsen, O., Long-term residential exposure to PM_2.5_, PM_10_, black carbon, NO_2_, and ozone and mortality in a Danish cohort. Environ Int 2019, 123, 265–272.

55. Kazemiparkouhi, F.; Eum, K. D.; Wang, B.; Manjourides, J.; Suh, H. H., Long-term ozone exposures and cause-specific mortality in a US Medicare cohort. J Expo Sci Environ Epidemiol 2020, 30, (4), 650–658.

56. Lim, C. C.; Hayes, R. B.; Ahn, J.; Shao, Y.; Silverman, D. T.; Jones, R. R.; Garcia, C.; Bell, M. L.; Thurston, G. D., Long-Term Exposure to Ozone and Cause-Specific Mortality Risk in the United States. Am J Respir Crit Care Med 2019, 200, (8), 1022–1031.

57. Paul, L. A.; Burnett, R. T.; Kwong, J. C.; Hystad, P.; van Donkelaar, A.; Bai, L.; Goldberg, M. S.; Lavigne, E.; Copes, R.; Martin, R. V., The impact of air pollution on the incidence of diabetes and survival among prevalent diabetes cases. Environ Int 2020, 134, 105333.

58. Shi, L.; Rosenberg, A.; Wang, Y.; Liu, P.; Danesh Yazdi, M.; Requia, W.; Steenland, K.; Chang, H.; Sarnat, J. A.; Wang, W.; Zhang, K.; Zhao, J.; Schwartz, J., Low-Concentration Air Pollution and Mortality in American Older Adults: A National Cohort Analysis (2001-2017). Environ Sci Technol 2021.

59. Strak, M.; Weinmayr, G.; Rodopoulou, S.; Chen, J.; de Hoogh, K.; Andersen, Z. J.; Atkinson, R.; Bauwelinck, M.; Bekkevold, T.; Bellander, T.; Boutron-Ruault, M. C.; Brandt, J.; Cesaroni, G.; Concin, H.; Fecht, D.; Forastiere, F.; Gulliver, J.; Hertel, O.; Hoffmann, B.; Hvidtfeldt, U. A.; Janssen, N. A. H.; Jockel, K. H.; Jorgensen, J. T.; Ketzel, M.; Klompmaker, J. O.; Lager, A.; Leander, K.; Liu, S.; Ljungman, P.; Magnusson, P. K. E.; Mehta, A. J.; Nagel, G.; Oftedal, B.; Pershagen, G.; Peters, A.; Raaschou-Nielsen, O.; Renzi, M.; Rizzuto, D.; van der Schouw, Y. T.; Schramm, S.; Severi, G.; Sigsgaard, T.; Sorensen, M.; Stafoggia, M.; Tjonneland, A.; Verschuren, W. M. M.; Vienneau, D.; Wolf, K.; Katsouyanni, K.; Brunekreef, B.; Hoek, G.; Samoli, E., Long term exposure to low level air pollution and mortality in eight European cohorts within the ELAPSE project: pooled analysis. BMJ 2021, 374.

60. Yazdi, M. D.; Wang, Y.; Di, Q.; Requia, W. J.; Wei, Y. G.; Shi, L. H.; Sabath, M. B.; Dominici, F.; Coull, B.; Evans, J. S.; Koutrakis, P.; Schwartz, J. D., Long-term effect of exposure to lower concentrations of air pollution on mortality among US Medicare participants and vulnerable subgroups: a doubly-robust approach. Lancet Planet Health 2021, 5, (10), E689–E697.

61. Bauwelinck, M.; Chen, J.; de Hoogh, K.; Katsouyanni, K.; Rodopoulou, S.; Samoli, E.; Andersen, Z. J.; Atkinson, R.; Casas, L.; Deboosere, P.; Demoury, C.; Janssen, N.; Klompmaker, J. O.; Lefebvre, W.; Mehta, A. J.; Nawrot, T. S.; Oftedal, B.; Renzi, M.; Stafoggia, M.; Strak, M.; Vandenheede, H.; Vanpoucke, C.; Van Nieuwenhuyse, A.; Vienneau, D.; Brunekreef, B.; Hoek, G., Variability in the association between long-term exposure to ambient air pollution and mortality by exposure assessment method and covariate adjustment: A census-based country-wide cohort study. Sci Total Environ 2022, 804, 150091.

62. Stafoggia, M.; Oftedal, B.; Chen, J.; Rodopoulou, S.; Renzi, M.; Atkinson, R. W.; Bauwelinck, M.; Klompmaker, J. O.; Mehta, A.; Vienneau, D., Long-term exposure to low ambient air pollution concentrations and mortality among 28 million people: results from seven large European cohorts within the ELAPSE project. Lancet Planet Health 2022, 6, (1), e9–e18.

63. Hvidtfeldt, U. A.; Geels, C.; Sørensen, M.; Ketzel, M.; Khan, J.; Tjønneland, A.; Christensen, J. H.; Brandt, J.; Raaschou-Nielsen, O., Long-term residential exposure to PM_2.5 _constituents and mortality in a Danish cohort. Environ Int 2019, 133, (Pt B), 105268.

64. Holtzman, M. J.; Cunningham, J. H.; Sheller, J. R.; Irsigler, G. B.; Nadel, J. A.; Boushey, H. A., Effect of ozone on bronchial reactivity in atopic and nonatopic subjects. Am Rev Respir Dis 1979, 120, (5), 1059–67.

65. Kodavanti, U. P.; Schladweiler, M. C.; Ledbetter, A. D.; Watkinson, W. P.; Campen, M. J.; Winsett, D. W.; Richards, J. R.; Crissman, K. M.; Hatch, G. E.; Costa, D. L., The spontaneously hypertensive rat as a model of human cardiovascular disease: Evidence of exacerbated cardiopulmonary injury and oxidative stress from inhaled emission particulate matter. Toxicol Appl Pharmacol 2000, 164, (3), 250–263.

66. Wang, M.; Sampson, P. D.; Sheppard, L. E.; Stein, J. H.; Vedal, S.; Kaufman, J. D., Long-Term Exposure to Ambient Ozone and Progression of Subclinical Arterial Disease: The Multi-Ethnic Study of Atherosclerosis and Air Pollution. Environ Health Perspect 2019, 127, (5), 57001.

67. Whyand, T.; Hurst, J. R.; Beckles, M.; Caplin, M. E., Pollution and respiratory disease: can diet or supplements help? A review. Respir Res 2018, 19, (1), 79.

68. EPA Integrated Science Assessment (ISA) for Particulate Matter; Washington, D.C., 2019.

69. World Health Organization, WHO global air quality guidelines: particulate matter (PM_2.5 _and PM_10_), ozone, nitrogen dioxide, sulfur dioxide and carbon monoxide: Executive summary. 2021.

70. Pope III, C. A.; Cohen, A. J.; Burnett, R. T., Cardiovascular disease and fine particulate matter: lessons and limitations of an integrated exposure-response approach. Circ Res 2018, 122, (12), 1645–1647.

71. Fantke, P.; McKone, T. E.; Tainio, M.; Jolliet, O.; Apte, J. S.; Stylianou, K. S.; Illner, N.; Marshall, J. D.; Choma, E. F.; Evans, J. S., Global effect factors for exposure to fine particulate matter. Environ Sci Technol 2019, 53, (12), 6855–6868.

72. Yin, P.; Brauer, M.; Cohen, A.; Burnett, R. T.; Liu, J.; Liu, Y.; Liang, R.; Wang, W.; Qi, J.; Wang, L.; Zhou, M., Long-term Fine Particulate Matter Exposure and Nonaccidental and Cause-specific Mortality in a Large National Cohort of Chinese Men. Environ Health Perspect 2017, 125, (11), 117002.

73. Malley, C. S.; Henze, D. K.; Kuylenstierna, J. C. I.; Vallack, H. W.; Davila, Y.; Anenberg, S. C.; Turner, M. C.; Ashmore, M. R., Updated Global Estimates of Respiratory Mortality in Adults >= 30 Years of Age Attributable to Long-Term Ozone Exposure. Environ Health Perspect 2017, 125, (8), 087021.

74. Anenberg, S. C.; Horowitz, L. W.; Tong, D. Q.; West, J. J., An estimate of the global burden of anthropogenic ozone and fine particulate matter on premature human mortality using atmospheric modeling. Environ Health Perspect 2010, 118, (9), 1189–1195.

75. Fann, N.; Lamson, A. D.; Anenberg, S. C.; Wesson, K.; Risley, D.; Hubbell, B. J., Estimating the national public health burden associated with exposure to ambient PM_2.5 _and ozone. Risk Anal 2012, 32, (1), 81–95.

76. Silva, R. A.; West, J. J.; Zhang, Y.; Anenberg, S. C.; Lamarque, J.-F.; Shindell, D. T.; Collins, W. J.; Dalsoren, S.; Faluvegi, G.; Folberth, G., Global premature mortality due to anthropogenic outdoor air pollution and the contribution of past climate change. Environ Res Lett 2013, 8, (3), 034005.

77. West, J. J.; Smith, S. J.; Silva, R. A.; Naik, V.; Zhang, Y.; Adelman, Z.; Fry, M. M.; Anenberg, S.; Horowitz, L. W.; Lamarque, J.-F., Co-benefits of mitigating global greenhouse gas emissions for future air quality and human health. Nat Clim Change 2013, 3, (10), 885–889.

78. Yu, P.; Guo, S.; Xu, R.; Ye, T.; Li, S.; Sim, M.; Abramson, M. J.; Guo, Y., Cohort studies of long-term exposure to outdoor particulate matter and risks of cancer: A systematic review and meta-analysis. The Innovation 2021, 100143.

79. Meng, X.; Liu, C.; Chen, R.; Sera, F.; Vicedo-Cabrera, A. M.; Milojevic, A.; Guo, Y.; Tong, S.; Coelho, M.; Saldiva, P. H. N.; Lavigne, E.; Correa, P. M.; Ortega, N. V.; Osorio, S.; Garcia Kysely, J.; Urban, A.; Orru, H.; Maasikmets, M.; Jaakkola, J. J. K.; Ryti, N.; Huber, V.; Schneider, A.; Katsouyanni, K.; Analitis, A.; Hashizume, M.; Honda, Y.; Ng, C. F. S.; Nunes, B.; Teixeira, J. P.; Holobaca, I. H.; Fratianni, S.; Kim, H.; Tobias, A.; Iniguez, C.; Forsberg, B.; Astrom, C.; Ragettli, M. S.; Guo, Y. L.; Pan, S. C.; Li, S.; Bell, M. L.; Zanobetti, A.; Schwartz, J.; Wu, T.; Gasparrini, A.; Kan, H., Short term associations of ambient nitrogen dioxide with daily total, cardiovascular, and respiratory mortality: multilocation analysis in 398 cities. BMJ 2021, 372, 534.

80. Yu, K.; Lv, J.; Qiu, G.; Yu, C.; Guo, Y.; Bian, Z.; Yang, L.; Chen, Y.; Wang, C.; Pan, A.; Liang, L.; Hu, F. B.; Chen, Z.; Li, L.; Wu, T.; China Kadoorie Biobank, S., Cooking fuels and risk of all-cause and cardiopulmonary mortality in urban China: a prospective cohort study. Lancet Glob Health 2020, 8, (3), e430–e439.

