## Supplementary Materials for "Long-term ozone exposure associated cause-specific mortality risks with adjusted metrics by cohort studies: A systematic review and meta-analysis" (from another version of this preprint: version 1, 2021-12-05)

*A total of 17 pages with 6 sections of texts, 6 tables and 7 figures.*

### Supplementary Texts

#### S1. Imputation procedures for exposure distribution

To reproduce the distribution, we first extracted the arithmetic means and standard deviations ($\sigma$) reported by literatures included for meta-analysis; if unavailable, we estimated the arithmetic means and standard deviations based on the reported descriptive statistics including median, first- and third-quartile, and all the other percentiles; and finally identified the corresponding distribution by Gaussian normal distribution assumption. Reported values were always treated as priority when multiple distribution parameters were given; and we also compared the reported values and our speculated values to check the credibility of our method.

During the process of exposure distribution reproducing, Gaussian normal distribution was presumed prior to log-normal distribution unless violated severely. The centric level, the arithmetic means and medians, were treated as exchangeable, but the arithmetic means were preferred. Theoretically, the minimum and maximum values of the distribution were not deductible, and thus $1^{st}$ and $99^{th}$ percentiles were used as proxies. Calculations for $\sigma$ from key percentiles followed: $75^{th}$%ile = mean + 0.6745 $\sigma$, $95^{th}$%ile = mean + 1.6449 $\sigma$, and $99^{th}$%ile = mean + 2.3263 $\sigma$. If IQRs were stated then IQR = 1.3490 $\sigma$; if the 5–$95^{th}$ percentile ranges were reported then range$_{5\text{-}95}$ = 3.2898 $\sigma$; if full minimum-maximum ranges were given then range = 4.6527 $\sigma$. If more than one distribution features were provided, IQRs were more preferred for $\sigma$ estimation for higher robustness.

The resampled distributions of $O_3$ exposure were listed in Table A for original reported metrics and in Table B for harmonised metric into 6mDMA8. By comparing the reported percentiles with distribution-based reproduced estimation values, reasonability of the distribution resampling was verified (Figure A) as the fitting $R^2$ = 0.84, RMSE = 2.78 ppbV. Theoretically, the minimum and maximum values of the distribution were not deductible, and thus $1^{st}$ and $99^{th}$ percentiles were used as surrogates.

**Table A. Originally reported and statistically reproduced long-term $O_3$ exposure distributions statistics for all meta-analysis included studies.** The originally reported long-term $O_3$ exposure levels from literatures were marked in **Bold** font, and the statistically reproduced exposure levels were denoted in brackets. All the exposure concentrations were uniformly scaled into ppbV.

| Study | Metric (ppbV) | Mean | STD | Min | 5% | 25% | Median | 75% | 95% | Max | IQR | Range |
|---|---|---|---|---|---|---|---|---|---|---|---|---|
| Abbey et al. 1999 | ADA24 | **26.1** | **7.7** | (8.3) | (13.5) | (21.0) | (26.1) | (31.3) | (38.7) | (43.9) | **12.0** (10.3) | **43.9** (35.6) |
| Lipfert et al. 2006 | ADMA1 | **84.2** | **10.2** | **38.5** (60.5) | (67.4) | (77.3) | (84.2) | (91.1) | (101) | **112** (108) | (13.8) | (73.5) |
| | ADA24 | **54.6** | **6.5** | **24.3** (39.5) | (43.9) | (50.2) | (54.6) | (59.0) | (65.3) | **72.1** (69.7) | (8.8) | (47.8) |
| Jerrett et al. 2009 | 6mDMA1 | | | | | | | | | | | |
| Krewski et al. 2009 | 6mDMA1 | (60.5)[1] | (15.2) | **33.3** | (35.5) | (50.3) | (60.5) | (70.7) | (85.5) | **104** | (20.5) | **70.7** |
| Smith et al. 2009 | 6mDMA1 | | | | | | | | | | | |
| Lipsett et al. 2011 | ADMA8 | **48.1** | **8.7** | **25.4** | (33.8) | (42.2) | (48.1) | (54.0) | (62.5) | **82.6** | **11.2** | **57.2** |
| Zanobetti et al. 2011 | 6mDMA8 | **46.0** | 5.2 | **26.6**[2] | (40.2) | **44.2** | **48.6** | **51.2** | (52.6) | **71.4** | **7.0** | **44.8** |
| Carey et al. 2013 | ADA24 | **26.4** | **1.2** | **22.7** (23.6) | (24.4) | (25.6) | (26.4) | (27.2) | (28.4) | **32.1** (29.2) | **1.5** (1.6) | **9.4** (5.6) |
| Jerrett et al. 2013 | ADMA8 | **50.4** | **14.6** | **17.1** (16.8) | **28.8** (26.8) | **36.8** (41.0) | **50.8** | **61.0** (60.6) | **74.2** (74.8) | **89.3** (84.8) | **24.2** (19.6) | **72.2** (68.0) |
| Bentayeb et al. 2015 | 6mDMA8 | **49.5** | **4.9** | (20.4) | (25.5) | (45.7) | **49.0** | (52.3) | (57.1) | (60.4) | **6.3** (6.6) | (40.0) |
| Crouse et al. 2015 | 6mDMA8 | **39.6** | (7.3) | **10.7** (22.0) | (26.9) | **34.3** (34.1) | **39.0** | **44.2** (43.9) | (51.1) | **60.0** (56.0) | **9.9** (9.8) | (49.3) |

| Study | Metric (ppbV) | Mean | STD | Min | 5% | 25% | Median | 75% | 95% | Max | IQR | Range |
|---|---|---|---|---|---|---|---|---|---|---|---|---|
| Tonne et al. 2016 | ADA24 | **20.6** | **2.0** | (15.9) | (17.3) | **19.3** (19.4) | **20.7** | **22.0** (22.0) | (24.0) | (25.4) | **2.7** | (9.5) |
| Turner et al. 2016 | ADMA8 | **38.2** | **4.0** | **26.7** (28.9) | (31.6) | (35.5) | (38.2) | (40.9) | (44.8) | **59.3** (47.5) | (5.4) | **32.6** (18.6) |
| Weichenthal et al. 2017 | 6mDMA8 | (38.2) | (6.6) | **1.0** (22.7) | **27.6** (27.2) | **33.7** | **38.1** | **42.6** | **50.5** (49.0) | **60.5** (53.5) | **9.0** | **59.5** (30.8) |
| Cakmak et al. 2018 | 6mDMA8 | **39.2** | **6.7** | **0.0** (23.6) | (28.2) | (34.7) | (39.2) | (43.7) | (50.2) | **58.8** (54.7) | (9.0) | **58.8** (31.1) |
| Di et al. 2018 | 6mDA24 | **46.3** | (6.0) | (32.4) | **36.3** | (42.3) | **46.3** | (50.3) | **55.9** | (60.2) | (8.0) | (27.7) |
| Hvidtfeldt et al. 2019 | ADA24 | (28.3) | (2.5) | (22.5) | **22.8** | (26.6) | **28.3** | (29.9) | **31.0** | (34.1) | (3.4) | (11.6) |
| Lim et al. 2019 | ADMA8 | **39.0** | **4.6** | **26.8** (28.3) | (31.4) | (35.9) | (39.0) | (42.1) | (46.6) | **56.3** (49.7) | (6.2) | **29.5** (21.4) |
| | 6mDMA8 | **46.2** | **7.6** | **29.5** (28.5) | (33.7) | (41.1) | (46.2) | (51.3) | (58.7) | **70.4** (63.9) | (10.3) | **40.9** (35.4) |
| Kazemiparkouhi et al. 2019 | 6mDMA1 | (55.0) | (7.4) | (37.8) | (42.8) | **50.0** | **55.0** | **60.0** | (67.2) | (72.2) | **10.0** | (34.5) |
| Paul et al. 2020 | 6mDMA8 | **46.9** | (4.7) | (35.9) | (39.1) | (43.7) | (46.9) | (50.1) | (54.7) | (57.9) | **6.4** | (22.0) |

§ Jerrett et al. 2009 did not report the arithmetic mean and standard deviation directly. The values were deducted by weighted averaging the centric concentrations of 4 exposure intervals on the populations given in Table 1 from the original literature.
⊥ Zanobetti et al. 2011 did not report the exposure distribution features directly. The quartiles were extracted from the legends provided in Figure 1 of the original literature.

**Table B. Statistically resampled distributions of $O_3$ exposure levels for each study included for meta-analysis.** The distribution features included arithmetic mean, standard deviation (STD), minimum, 5th, 25th, 50th (median), 75th, and 95th percentile, maximum, inter-quartile range (IQR), and full range, based on $O_3$ exposure concentrations scaled by 6mDMA8 metric in ppbV. Values in **Bold** font represented the statistics reported by literature, while the rest indicated resampled values.

| Study | Mean | STD | Min | 5% | 25% | Median | 75% | 95% | Max | IQR | Range |
|---|---|---|---|---|---|---|---|---|---|---|---|
| **Abbey et al. 1999** | **50.4** | **14.9** | 16.1 | 26.1 | 40.6 | 50.4 | 60.5 | 74.8 | 84.9 | **23.2** | **84.9** |
| **Lipfert et al. 2006** | **80.1** | **9.7** | **36.6** | 64.2 | 73.5 | 80.1 | 86.7 | 96.1 | **106.6** | 13.2 | 69.9 |
| **Jerrett et al. 2009** | | | | | | | | | | | |
| **Krewski et al. 2009** | 50.1 | 12.6 | **27.5** | 30.0 | 41.6 | 50.1 | 58.5 | 70.7 | **86.1** | 17.0 | **58.5** |
| **Smith et al. 2009** | | | | | | | | | | | |
| **Lipsett et al. 2011** | **55.6** | **10.1** | **29.4** | 39.1 | 48.8 | 55.6 | 62.4 | 72.3 | **95.5** | **12.9** | **66.1** |
| **Zanobetti et al. 2011** | **45.9** | 5.2 | **26.6** | **40.1** | **44.0** | **48.4** | **51.1** | **52.5** | **71.2** | **6.9** | **44.7** |
| **Carey et al. 2013** | **51.0** | **2.3** | **43.8** | 47.2 | 49.5 | 51.0 | 52.6 | 54.9 | **62.0** | **2.9** | **18.1** |
| **Jerrett et al. 2013** | **58.3** | **16.9** | **19.8** | **33.3** | **42.5** | **58.7** | **70.5** | **85.8** | **103.2** | **28.0** | **83.5** |
| **Bentayeb et al. 2015** | **49.4** | **4.9** | 20.3 | 25.4 | 45.5 | **48.9** | 52.1 | 57.0 | 60.2 | **6.2** | 39.9 |
| **Crouse et al. 2015** | **39.5** | 7.3 | **10.7** | 26.8 | **34.2** | **39.0** | **44.0** | 51.0 | **59.9** | **9.8** | 49.1 |
| **Tonne et al. 2016** | **39.8** | **3.8** | 30.7 | 33.4 | **37.3** | **40.0** | **42.5** | 46.4 | 49.0 | **5.2** | 18.4 |
| **Turner et al. 2016** | **44.2** | **4.6** | **30.1** | 36.5 | 41.0 | 44.2 | 47.3 | 51.8 | **68.6** | 6.2 | **37.7** |
| **Weichenthal et al. 2017** | 38.1 | 6.6 | **1.0** | **27.5** | **33.6** | **38.0** | **42.5** | **50.4** | **60.3** | **9.0** | **59.3** |
| **Cakmak et al. 2018** | **39.1** | **6.7** | **0.0** | 28.1 | 34.6 | 39.1 | 43.6 | 50.1 | **58.6** | 9.0 | **58.6** |
| **Di et al. 2018** | **77.1** | 9.9 | 54.0 | **60.5** | 70.5 | **77.1** | 83.8 | **93.2** | 100.2 | 13.3 | 46.1 |
| **Hvidtfeldt et al. 2019** | 54.7 | 4.9 | 43.5 | **44.0** | 51.3 | **54.7** | 57.8 | **59.9** | 65.9 | 6.6 | 22.4 |
| **Kazemiparkouhi et al. 2019** | **45.1** | **5.3** | **31.0** | 36.3 | 41.5 | 45.1 | 48.7 | 53.9 | **65.1** | 7.2 | **34.1** |
| **Lim et al. 2019** | 45.5 | 6.1 | 31.3 | 35.4 | **41.4** | **45.5** | **49.7** | 55.6 | 59.8 | **8.3** | 28.6 |
| **Paul et al. 2020** | **46.8** | 4.7 | 35.8 | 39.0 | 43.6 | 46.8 | 49.9 | 54.6 | 57.8 | **6.4** | 22.0 |

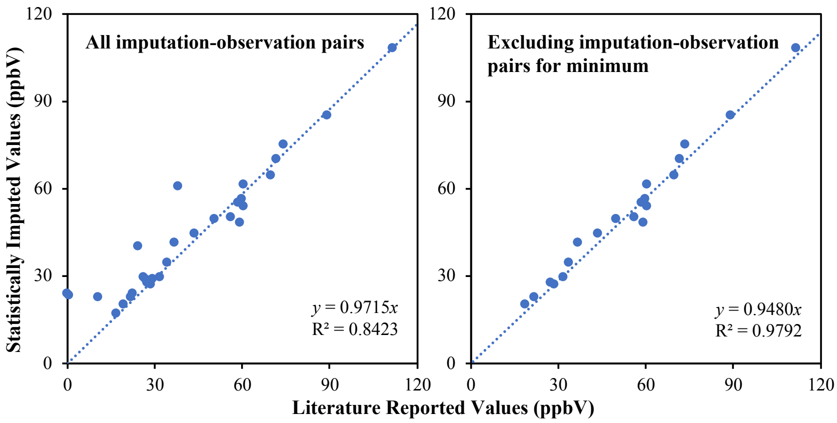

**Figure A. Accuracy evaluation of imputed distribution statistics for 20 studies included for systematic review and meta-analysis.** The left panel indicated pair-wise accuracies for all distribution statistics including minimum, 5th, 25th, 75th, 95th percentile, and maximum, and the right panel excluded the minimum. Non-intercept linear models were applied to estimate the coefficients of determination ($R^2$).

#### S2. Enhanced integrated exposure-response curve-fitting

Exposure-response quantitative relationships were regarded as an additive credit to risk association estimations, as in many cases the adverse health outcomes would respond to exposures in non-linear patterns.[1] We developed the exposure-response non-linear relationships following the integrated exposure-response (IER) curve-fitting approach,[2] based on the mathematical function as:

$$RR(x) = \begin{cases} 1, & x < x_{cf} \\ 1 + \alpha\left(1 - e^{-\gamma (x - x_{cf})^{\delta}}\right), & x \geq x_{cf} \end{cases}$$

where the subscript *cf* refers to a counterfactual concentration below which no additional health risks are assumed, exchangeable with the concept of "threshold" or "low concentration cut-off" (LCC).[3]

However, this method required more sufficient sampling of the exposure levels, since small number of studies could only provide limited exposure values to be fitted, which would impair the stability of the curve estimations. In addition, simply drawing an averaged exposure level from a high-quality environmental health study would lose exposure variability information. In this event, we intended to take better advantage of the variabilities in exposure levels by statistically imputing the exposure distributions for each study from the provided statistics mainly including the mean, standard deviation, and percentiles, rather than just using a simple centric averaged exposure concentration, for curve fitting.

To enhance the exposure-response trend construction, we first set the exposure interval nodes by every 5-ppbV ranging between the minimum and maximum exposures reported from all included literatures. In the second step, we selected the studies of which the exposure levels contained each specific node. Thirdly, the counterfactual concentrations were defined as the lowest 5th percentile following a previous research,[3] based on which the cumulative risks at the prescribed nodes were calculated for each study. Fourthly, the individual cumulative risks were pooled into a synthesised risk with 95% UI for each exposure node. As for the fifth step, we generated 10,000 groups of concentration-risk pairs by Monte Carlo random draws according to the pooled risks with 95% UIs, and for each draw the counterfactual concentration was randomly sampled from a uniform distribution between the minimum and 5th lowest percentile concentrations, based on which the draw-specific curve-shape parameters (α, γ, and δ) were estimated. At last, we summarised the distribution patterns of the parameters from all randomised draws, and reproduced the concentration-response curve by another 10,000-time Monte Carlo simulation to characterise the uncertainties. The Monte Carlo simulations were performed in MathWorks® MATLAB 2016b, and the curve fittings were finished in Python 3.8.5 by package Scipy 1.5.2.

#### S3. Demonstrative procedures of enhanced exposure-response trend curve-fitting

Here is a detailed elaborative demonstration for COPD mortality risk curve-fitting by 6mDMA8 metric.

First, identify studies covering each exposure concentration interval level, hereafter called as "*nodes*". According to the resampled exposure distribution, 25.4 ppbV was the lowest $5^{th}$%ile concentration reported (Table B), and thence 30 ppbV could be set as the starting node, followed with every 5-ppbV increment as 35, 40, ..., 80 ppbV. For an instance, treating the lowest $5^{th}$ percentile as the effective lowest exposure level for each study, 5 studies as Abbey et al. (1999), Bentayeb et al. (2015), Crouse et al. (2015), Weichenthal et al. (2017) and Cakmak et al. (2018) covered the minimum node as 30 ppbV by 6mDMA8 metric (Table B), and likewise for the rest nodes. Among the 5 studies identified, only 2 studies as Crouse et al. (2015) and Cakmak et al. (2018) would be used when exploring COPD mortality risks since the rest 3 did not involve COPD studies.

Second, for each exposure node, calculate the study-specific cumulative RRs. Taking Crouse et al. (2015) as an example from the 2 studies covering 30-ppbV exposure, the $5^{th}$ percentile exposure, 26.8 ppbV by 6mDMA8 metric, was set as the threshold, and hence the cumulative RR for COPD mortality at 30 ppbV exposure should be $e^{(ln0.959\times\frac{30-26.8}{10})}$ = 0.987, together with its 95% CI: 0.975-0.999. Cumulative RRs for the rest studies and other cause-specific mortalities were calculated in the same way.

Third, pool the study-specific individual risks into overall cumulative RRs at each note by meta-regression within each mortality cause. For COPD mortality risks at 30-ppbV exposure, the cumulative RR by Crouse et al. (2015) was 0.987 (95% CI: 0.975–0.999), and by Cakmak et al. (2018) was 1.000 (95% CI: 0.998–1.002), pooling from which the overall cumulative RR was 0.997 (95% CI:0.990–1.003). The rest intervals followed the same procedure, as summarised in Table C, which could then be applied onto other mortality causes as listed in Table D.

Fourth, for each exposure node, draw a random cumulative risk value based on the normal distribution presumption; and generate a random value between 0 and the lowest $5^{th}$ percentile across all included studies as the overall threshold level. Then, fit the drawn "node-risk" combinations by the prescribed mathematical formula as presented in Supplementary Text S1, from which three parameters α, δ, and γ could be estimated.

Fifth, duplicate the fourth procedure for 10,000 times with Monte Carlo simulation, and acquire 10,000 groups of the estimated parameter sets, based on which the arithmetic mean, $2.5^{th}$ and $97.5^{th}$ percentiles were generated.

Finally, the exposure-response trend curves were reproduced by another Monte Carlo simulation using the estimated 3 parameters together with the uniformly distribution presumed threshold level (between 0 and the cross-study lowest $5^{th}$ percentile level) to calculate the cumulative RRs through the full range of exposures. The integrated exposure-response curves were plotted in Figure B.

**Table C. Cumulative relative risks of chronic obstructive pulmonary disease (COPD) mortality by reproduced $O_3$ exposure distribution.**

| Study | 30 ppbV | 35 ppbV | 40 ppbV | 45 ppbV | 50 ppbV | 55 ppbV | 60 ppbV |
|---|---|---|---|---|---|---|---|
| Zanobetti et al. 2011 | | 1.23 (1.13, 1.30) | 1.33 (1.18, 1.43) | 1.43 (1.23, 1.58) | 1.55 (1.29, 1.75) | 1.68 (1.35, 1.93) | 1.81 (1.41, 2.14) |
| Crouse et al. 2015 | 0.99 (0.98, 1.00) | 0.96 (0.83, 0.99) | 0.93 (0.88, 0.99) | 0.90 (0.83, 0.99) | 0.87 (0.78, 0.99) | | |
| Turner et al. 2016 | | 1.19 (1.11, 1.28) | 1.27 (1.15, 1.41) | 1.35 (1.19, 1.55) | 1.44 (1.24, 1.71) | 1.54 (1.29, 1.88) | |
| Cakmak et al. 2018 | 1.00 (0.99, 1.00) | 1.00 (0.96, 1.05) | 1.00 (0.94, 1.06) | 1.00 (0.92, 1.08) | 1.00 (0.91, 1.10) | | |
| Kazemiparkouhi et al. 2019 | | 1.16 (1.15, 1.17) | 1.22 (1.21, 1.24) | 1.30 (1.28, 1.32) | 1.37 (1.35, 1.40) | | |
| Lim et al. 2019 | | 1.09 (1.02, 1.19) | 1.13 (1.02, 1.27) | 1.17 (1.03, 1.35) | 1.21 (1.03, 1.44) | 1.25 (1.04, 1.54) | |
| **Heterogeneity $I^2$** | 65.3% | 94.7% | 94.7% | 94.7% | 94.7% | 58.7% | - |
| **Pooled RRs** | **1.00 (0.99, 1.00)** | **1.09 (1.01, 1.19)** | **1.13 (1.01, 1.27)** | **1.17 (1.02, 1.35)** | **1.21 (1.02, 1.44)** | **1.48 (1.25, 1.76)** | **1.81 (1.41, 2.14)** |

**Table D.** **Relative risks of all-cause, respiratory, cardiovascular and congestive heart failure mortality by distribution-reproduced $O_3$ exposures.**

| Concentrations | RR | LB | UB | $I^2$ |
|---|---|---|---|---|
| **All-cause Mortality** | | | | |
| 30 ppbV | 1.003 | 1.002 | 1.003 | 93.2% |
| 35 ppbV | 1.008 | 1.007 | 1.009 | 93.8% |
| 40 ppbV | 1.006 | 1.004 | 1.008 | 93.2% |
| 45 ppbV | 1.008 | 1.005 | 1.010 | 93.7% |
| 50 ppbV | 1.009 | 1.006 | 1.012 | 93.5% |
| 55 ppbV | 1.021 | 1.019 | 1.023 | 85.6% |
| 60 ppbV | 1.025 | 1.022 | 1.027 | 84.1% |
| 65 ppbV | 1.028 | 1.025 | 1.030 | 84.1% |
| 70 ppbV | 1.039 | 1.035 | 1.043 | 84.1% |
| 75 ppbV | 1.040 | 1.036 | 1.043 | 84.7% |
| 80 ppbV | 1.040 | 1.036 | 1.043 | 84.7% |
| **Respiratory Mortality** | | | | |
| 30 ppbV | 1.088 | 1.079 | 1.096 | 94.5% |
| 35 ppbV | 1.090 | 1.082 | 1.098 | 97.0% |
| 40 ppbV | 1.094 | 1.089 | 1.100 | 94.3% |
| 45 ppbV | 1.094 | 1.089 | 1.100 | 96.0% |
| 50 ppbV | 1.096 | 1.090 | 1.102 | 96.1% |
| 55 ppbV | 1.099 | 1.093 | 1.106 | 93.6% |
| 60 ppbV | 1.098 | 1.094 | 1.101 | 51.7% |
| 65 ppbV | 1.098 | 1.094 | 1.101 | 48.7% |
| 70 ppbV | 1.098 | 1.094 | 1.101 | 46.4% |
| 75 ppbV | 1.093 | 1.067 | 1.118 | 5.5% |
| **Cardiovascular Mortality** | | | | |
| 30 ppbV | 1.028 | 0.984 | 1.071 | 98.5% |
| 35 ppbV | 1.042 | 1.007 | 1.079 | 98.1% |
| 40 ppbV | 1.048 | 1.003 | 1.096 | 97.9% |
| 45 ppbV | 1.062 | 1.003 | 1.125 | 97.9% |
| 50 ppbV | 1.077 | 1.003 | 1.154 | 97.9% |
| 55 ppbV | 1.010 | 0.965 | 1.057 | 83.4% |
| 60 ppbV | 1.008 | 0.955 | 1.063 | 79.6% |
| 65 ppbV | 1.009 | 0.949 | 1.072 | 79.6% |
| 70 ppbV | 1.010 | 0.944 | 1.081 | 79.6% |
| **Congestive Heart Failure Mortality** | | | | |
| 35 ppbV | 1.095 | 1.051 | 1.142 | 84.9% |
| 40 ppbV | 1.134 | 1.071 | 1.201 | 84.9% |
| 45 ppbV | 1.174 | 1.091 | 1.264 | 84.9% |
| 50 ppbV | 1.216 | 1.112 | 1.330 | 84.9% |
| 55 ppbV | 1.234 | 1.030 | 1.478 | 87.5% |
| 60 ppbV | 1.254 | 1.122 | 1.349 | - |

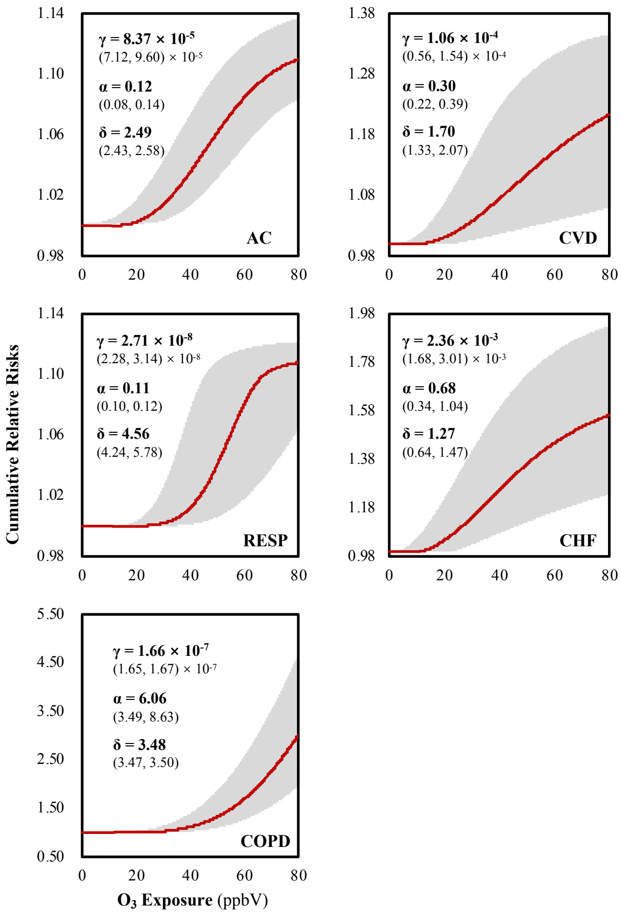

**Figure B.** **Estimated exposure-response curves for mortality risks from all-cause (AC), all respiratory diseases (RESP), chronic obstructive pulmonary disease (COPD), all cardiovascular diseases (CVD), and congestive heart failure (CHF).** The trend curves were plotted by centric cumulative relative risk values with 95% uncertainty intervals (95% UI). Estimates of 3 parameters were shown with 95% confidence intervals (95% CI).

Mathematically speaking, the cumulative risks will flatten at $1 + \alpha$, which indicates the anticipated highest risks. For all-cause $O_3$-associated mortality, the cumulative risk would saturate at around RR = 1.12 by 6mDMA8 metric of $O_3$ exposure; respiratory mortality at RR = 1.11; cardiovascular mortality at RR = 1.30; and congestive heart failure mortality at RR = 1.68; while COPD mortality risks did not manifest level-off tendency in observed exposure range, as the cumulative risks kept speedily climbing till 80 ppbV.

Large uncertainties were observed in the Monte-Carlo fitted exposure-response curves, which should mainly be ascribed to the limited number of large-scale cohort-based epidemiological studies. We therefore suggest researchers critically scrutinise this methodology and cautiously use our results.

#### S4. Undefined metric imputation

For Carey et al. (2013), the $O_3$ exposure metric was not stated clearly, but exposure concentrations were reported as 26.4 ± 1.2 ranging between 22.7–32.1 ppbV. Since the cohort study was conducted in UK, we checked the corresponding concentrations spatiotemporal closely from TOAR observations, and we found by

ADA24 metric the average $O_3$ level was 25.7 ± 8.3 ppbV, with IQR 19.8–31.3 ppbV. We thus deduced the metric should be ADA24.

For Jerrett et al. (2009), not any $O_3$ exposure distributions were reported, but only the minimum-maximum range was given as 33.3–104.0 ppbV. We calculated the average level by our own, as weighting the population with interval centric point: 43.2 ppbV by 126 206, 55.3 ppbV by 95 740, 59.95 ppbV by 106 545, and 83.25 ppbV by 120 359, to obtain the estimated average as 60.5 ppbV. According to the US observations by TOAR, the average levels were around 62.8 ppbV by warm-season DMA1 and 50.9 by full-year DMA1, the former of which was closer to the literature-generated value, 60.5 ppbV, and we thus inferred the exposure metric should be 6mDMA1 more reasonably.

Jerrett et al. (2013) did not report which metric they used, but detailed distribution patterns were provided. Based on TOAR observations by ADMA8 in US, $O_3$ concentrations were 54.1 ± 16.1 (IQR: 42.4–65.5, range: 17.6–102.5) ppbV, closer to the reported values as 50.4 ± 14.6 (IQR: 36.8–61.0, range: 17.1–89.3) ppbV than 6mDMA8 which was of average levels as 63.1 ± 18.2 ppbV.

#### S5. Interpretation and procedure of cross-metric linear conversion

For direct cross-metric linear conversion, taking conversion from ADA24 to ADMA8 as an interpretation example, the prediction could be approximated by ADMA8 = 1.67 × ADA24 ($R^2$ = 0.97, **Figure 2**), which means each 10-ppbV change by ADA24 metric should correspond to 16.7-ppbV change by ADMA8 metric, and likewise for the rest conversions. The conversion factors deviant from 1 revealed that unit changes in $O_3$ concentrations by different metrics could be of discrepant realistic implications, and hence unifications of $O_3$ exposure metrics should be necessary prior to meta-analysis.

For cross-metric relative risk conversion, taking Smith et al. (2009) as an example, the originally reported all respiratory mortality HR = 1.0057 (95% CI: 1.0020–1.0094) by every 1-μg/$m^3$ incremental $O_3$ exposure by 6mMDA1, equally HR = 1.0585 (95% CI: 1.0202–1.0981) by every 10-μg/$m^3$ (equal to 10/1.96 = 5.10-ppbV) incremental $O_3$ exposure. Converting μg/$m^3$ into ppbV, every 10-ppbV increasing $O_3$ exposure by 6mMDA1 should correspond to HR = 1.1179 (95% CI: 1.0400–1.2013). Following the metric-conversion coefficient estimation, 10-ppbV by 6mMDA1 was identical to 8.31-ppbV by 6mDMA8 (**Figure 2**), and thus HR should be scaled to 1.1435 (95% CI: 1.0483–1.2470) for every 10-ppbV incremental exposure by 6mDMA8.

#### S6. Detailed study assessment

For cohort study quality assessment (**Table S4**), 6 studies were ranked as "Good" and the rest 14 as "Fair" in summary. All studies well met 10 out of 14 assessment items, while 9 studies did not clarify their inclusion and exclusion criteria;[4-12] 2 re-analysis study reports did not clearly state the $O_3$ exposures;[5, 13] 2 studies were of relatively insufficient follow-up durations, e.g. less than 5 years, to observe the outcomes;[7, 11] and 10 studies were of methodological deficiencies in identifying the exposures,[4-7, 12-17] most of which were studies published before 2013 when data assimilation techniques were not adequately mature to fuse observations with other full spatial coverage products such as satellite-based remote sensing and atmospheric mechanistic simulations. The satisfactory assessment results indicated that the overall risks of bias were reasonably low, laying the reliable foundation for further meta-analyses.

For the epidemiological evidence credibility assessment (**Table S5**), the overall GRADE judgements for all-cause, respiratory, and lung cancer mortality risks were "High", while for cerebrovascular mortality risk was "Low", and the rest cause-specific mortality risks were all "Moderate". Due to inconsistency in centric risk effects, 8 out of 9 cause-specific mortality risk assessments were downgraded, except for the congestive heart failure induced mortality risk. There were 4 studies examining the exposure-response trends,[10, 15, 18, 19] which upgraded the ratings of all-cause, respiratory and cardiovascular mortality risks accordingly. Cakmak et al. (2018) found confounding effects towards null hypothesis that adjusting the confounders had increased the positive mortality,[20] which upgraded the GRADE rating of all-cause, ischaemic heart disease and lung cancer mortality risks associated with long-term $O_3$ exposure. Due to limited studies screened out for meta-analysis, GRADE ratings were not conducted on long-term $O_3$ exposure-associated ischaemic stroke or pneumonia mortality risks.

### Supplementary Tables and Figures

**Table S1 Searching strategy.**

| | |
|---|---|
| #21 | #17 AND #18 AND #19 AND #20 |
| #20 | #14 OR #15 OR #16 |
| #19 | #11 OR #12 OR #13 |
| #18 | #3 OR #4 OR #5 OR #6 OR #7 OR #8 OR #9 OR #10 |
| #17 | #1 OR #2 |
| #16 | (ozone OR O3 OR "O 3" OR "O(3)" OR "O-3" OR "air pollution").TW. |
| #15 | (ozone OR O3 OR "O 3" OR "O(3)" OR "O-3" OR "air pollution").NM. |
| #14 | (ozone OR "air pollution").SH. |
| #13 | (long-term OR "long term").TW. |
| #12 | (cohort OR Cox OR hazard OR HR OR risk OR prospective OR retrospective).TW. |
| #11 | (cohort studies).SH. |
| #10 | (lung cancer).TW. |
| #9 | ("cerebrovascular disease" OR stroke).TW. |
| #8 | ("congestive heart failure" OR CHF).TW. |
| #7 | ("ischemic heart disease" OR "ischaemic heart disease" OR IHD).TW. |
| #6 | (cardiovascular OR CVD).TW. |
| #5 | (respiratory OR "chronic obstructive pulmonary disease" OR COPD OR pneumonia).TW. |
| #4 | (cardiopulmonary disease).TW. |
| #3 | (all-cause OR "all cause").TW. |
| #2 | (mortality OR death OR "premature death").TW. |
| #1 | (mortality OR death OR "premature death").SH. |

Search timeline: 1 September 2015 to 15 July 2021

SH: MeSH subject headings. NM: Name of substance word. TW: Text word.

**Table S2 Quality assessment tool for observational cohort and cross-sectional studies.**

A. Was the research question or objective in this paper clearly stated?

B. Was the study population clearly specified and defined?

C. Was the participation rate of eligible persons at least 50%?

D. Were all the subjects selected or recruited from the same or similar populations (including the same time period)? Were inclusion and exclusion criteria for being in the study prespecified and applied uniformly to all participants?

E. Was a sample size justification, power description, or variance and effect estimates provided?

F. For the analyses in this paper, were the exposure(s) of interest measured prior to the outcome(s) being measured?

G. Was the timeframe sufficient so that one could reasonably expect to see an association between exposure and outcome if it existed?

H. For exposures that can vary in amount or level, did the study examine different levels of the exposure as related to the outcome (e.g., categories of exposure, or exposure measured as continuous variable)?

I. Were the exposure measures (independent variables) clearly defined, valid, reliable, and implemented consistently across all study participants?

J. Was the exposure(s) assessed more than once over time?

K. Were the outcome measures (dependent variables) clearly defined, valid, reliable, and implemented consistently across all study participants?

L. Were the outcome assessors blinded to the exposure status of participants?

M. Was loss to follow-up after baseline 20% or less?

N. Were key potential confounding variables measured and adjusted statistically for their impact on the relationship between exposure(s) and outcome(s)?

Source: https://www.nhlbi.nih.gov/health-topics/study-quality-assessment-tools.

**Table S3 Harmonised relative risks of 8 causes of mortality by 10-ppbV incremental $O_3$ exposures in 6mDMA8 metric.**

| Study | RR | LB | UB |
|---|---|---|---|
| **All-cause (AC)** | | | |
| Abbey et al. 1999 (male cohort) | 1.058 | 0.966 | 1.159 |
| Abbey et al. 1999 (female cohort) | 0.966 | 0.898 | 1.040 |
| Lipfert et al. 2006b | 1.030 | 1.011 | 1.049 |
| Jerrett et al. 2009 | 0.988 | 0.979 | 0.995 |
| Krewski et al. 2009 | 1.022 | 1.011 | 1.033 |
| Smith et al. 2009 | 1.004 | 0.982 | 1.031 |
| Lipsett et al. 2011 | 0.994 | 0.987 | 1.002 |
| Carey et al. 2013 | 0.880 | 0.798 | 0.939 |
| Jerrett et al. 2013 | 1.000 | 0.992 | 1.007 |
| Bentayeb et al. 2015 | 0.829 | 0.668 | 1.029 |
| Crouse et al. 2015 | 1.017 | 1.010 | 1.025 |
| Tonne et al. 2016 | 0.965 | 0.847 | 1.090 |
| Turner et al. 2016 | 1.016 | 1.008 | 1.032 |

| Study | RR | LB | UB |
|---|---|---|---|
| Weichenthal et al. 2017 | 1.054 | 1.044 | 1.062 |
| Cakmak et al. 2018 | 1.073 | 1.018 | 1.129 |
| Di et al. 2018 | 1.006 | 1.006 | 1.006 |
| Hvidtfeldt et al. 2019 | 0.954 | 0.915 | 1.000 |
| Kazemiparkouhi et al. 2019 | 1.002 | 1.001 | 1.003 |
| Lim et al. 2019 | 1.000 | 0.990 | 1.010 |
| **Chronic obstructive pulmonary disease (COPD)** | | | |
| Zanobetti et al. 2011 | 1.133 | 1.075 | 1.172 |
| Crouse et al. 2015 | 0.962 | 0.930 | 0.996 |
| Turner et al. 2016 | 1.110 | 1.063 | 1.164 |
| Cakmak et al. 2018 | 1.000 | 0.972 | 1.028 |
| Kazemiparkouhi et al. 2019 | 1.094 | 1.088 | 1.099 |
| Lim et al. 2019 | 1.055 | 1.010 | 1.110 |

**Table S3** (cont'd) **Harmonised relative risks of 8 causes of mortality by 10-ppbV incremental $O_3$ exposures in 6mDMA8 metric.**

| Study | RR | LB | UB |
|---|---|---|---|
| **Respiratory diseases (RESP)** | | | |
| Abbey et al. 1999 (male cohort) | 1.078 | 0.898 | 1.290 |
| Abbey et al. 1999 (female cohort) | 1.033 | 0.877 | 1.219 |
| Jerrett et al. 2009 | 1.044 | 1.014 | 1.075 |
| Smith et al. 2009 | 1.132 | 1.044 | 1.226 |
| Lipsett et al. 2011 | 1.018 | 0.994 | 1.040 |
| Carey et al. 2013 | 0.798 | 0.720 | 0.880 |
| Jerrett et al. 2013 | 1.003 | 0.980 | 1.028 |
| Bentayeb et al. 2015 | 0.956 | 0.580 | 1.606 |
| Crouse et al. 2015 | 0.982 | 0.956 | 1.007 |
| Turner et al. 2016 | 1.094 | 1.063 | 1.125 |
| Weichenthal et al. 2017 | 1.038 | 1.010 | 1.065 |
| Hvidtfeldt et al. 2019 | 0.972 | 0.897 | 1.047 |
| Kazemiparkouhi et al. 2019 | 1.036 | 1.033 | 1.041 |
| Lim et al. 2019 | 1.036 | 1.000 | 1.073 |
| **Cardiovascular diseases (CVD)** | | | |
| Jerrett et al. 2009 | 0.981 | 0.968 | 0.994 |
| Smith et al. 2009 | 1.049 | 1.014 | 1.105 |
| Lipsett et al. 2011 | 1.004 | 0.991 | 1.014 |
| Jerrett et al. 2013 | 1.009 | 0.998 | 1.021 |
| Bentayeb et al. 2015 | 0.842 | 0.427 | 1.657 |
| Crouse et al. 2015 | 1.037 | 1.023 | 1.051 |
| Turner et al. 2016 | 1.024 | 1.008 | 1.040 |
| Weichenthal et al. 2017 | 1.148 | 1.132 | 1.163 |
| Hvidtfeldt et al. 2019 | 0.941 | 0.910 | 0.981 |
| Kazemiparkouhi et al. 2019 | 0.997 | 0.994 | 0.999 |
| Lim et al. 2019 | 1.018 | 0.990 | 1.028 |
| Paul et al. 2020 | 1.097 | 1.071 | 1.122 |
| **Congestive heart failure (CHF)** | | | |
| Zanobetti et al. 2011 | 1.056 | 1.029 | 1.075 |
| Turner et al. 2016 | 1.118 | 1.079 | 1.156 |
| Kazemiparkouhi et al. 2019 | 1.080 | 1.070 | 1.089 |
| Lim et al. 2019 | 1.010 | 0.972 | 1.046 |

| Study | RR | LB | UB |
|---|---|---|---|
| **Ischaemic heart diseases (IHD)** | | | |
| Jerrett et al. 2009 | 0.970 | 0.954 | 0.986 |
| Krewski et al. 2009 | 1.011 | 0.989 | 1.022 |
| Lipsett et al. 2011 | 1.018 | 1.002 | 1.036 |
| Jerrett et al. 2013 | 1.020 | 1.004 | 1.036 |
| Crouse et al. 2015 | 1.060 | 1.044 | 1.077 |
| Turner et al. 2016 | 0.984 | 0.960 | 1.000 |
| Cakmak et al. 2018 | 1.110 | 1.092 | 1.119 |
| Kazemiparkouhi et al. 2019 | 0.995 | 0.992 | 0.999 |
| Lim et al. 2019 | 1.028 | 1.000 | 1.055 |
| **Cerebrovascular diseases (CEVD)** | | | |
| Lipsett et al. 2011 | 0.998 | 0.974 | 1.020 |
| Crouse et al. 2015 | 1.022 | 0.993 | 1.053 |
| Turner et al. 2016 | 1.024 | 0.984 | 1.055 |
| Kazemiparkouhi et al. 2019 | 0.986 | 0.980 | 0.990 |
| Lim et al. 2019 | 0.926 | 0.870 | 0.982 |
| **Lung cancer (LC)** | | | |
| Abbey et al. 1999 (male cohort) | 1.634 | 0.994 | 2.682 |
| Abbey et al. 1999 (female cohort) | 0.841 | 0.518 | 1.371 |
| Krewski et al. 2009 | 0.989 | 0.956 | 1.022 |
| Lipsett et al. 2011 | 0.990 | 0.960 | 1.020 |
| Carey et al. 2013 | 0.825 | 0.720 | 0.939 |
| Jerrett et al. 2013 | 0.971 | 0.944 | 0.998 |
| Crouse et al. 2015 | 0.974 | 0.951 | 0.997 |
| Turner et al. 2016 | 0.968 | 0.928 | 1.000 |
| Cakmak et al. 2018 | 1.036 | 0.972 | 1.110 |
| Kazemiparkouhi et al. 2019 | 1.017 | 1.012 | 1.023 |
| Lim et al. 2019 | 0.982 | 0.954 | 1.000 |

**Table S4 Quality assessment of 20 included cohort studies for meta-analysis.**

| Study | A | B | C | D | E | F | G | H | I | J | K | L | M | N | Score |
|---|---|---|---|---|---|---|---|---|---|---|---|---|---|---|---|
| Abbey et al. 1999 | √ | √ | √ | √ | √ | √ | √ | √ | | √ | √ | √ | √ | √ | Fair |
| Lipfert et al. 2006 | √ | √ | √ | | √ | √ | √ | √ | | √ | √ | √ | √ | √ | Fair |
| Jerrett et al. 2009 | √ | √ | √ | √ | √ | √ | √ | √ | | √ | √ | √ | √ | √ | Fair |
| Krewski et al. 2009 | √ | √ | √ | √ | √ | | √ | √ | | √ | √ | √ | √ | √ | Fair |
| Smith et al. 2009 | √ | √ | √ | | √ | | √ | √ | | √ | √ | √ | √ | √ | Fair |
| Lipsett et al. 2011 | √ | √ | √ | | √ | √ | √ | √ | | √ | √ | √ | √ | √ | Fair |
| Zanobetti et al. 2011 | √ | √ | √ | √ | √ | √ | √ | √ | | √ | √ | √ | √ | √ | Fair |
| Carey et al. 2013 | √ | √ | √ | | √ | √ | | √ | | √ | √ | √ | √ | √ | Fair |
| Jerrett et al. 2013 | √ | √ | √ | √ | √ | √ | √ | √ | | √ | √ | √ | √ | √ | Fair |
| Bentayeb et al. 2015 | √ | √ | √ | | √ | √ | √ | √ | √ | √ | √ | √ | √ | √ | Fair |
| Crouse et al. 2015 | √ | √ | √ | | √ | √ | √ | √ | √ | √ | √ | √ | √ | √ | Fair |
| Tonne et al. 2016 | √ | √ | √ | √ | √ | √ | √ | √ | √ | √ | √ | √ | √ | √ | Good |
| Turner et al. 2016 | √ | √ | √ | √ | √ | √ | √ | √ | √ | √ | √ | √ | √ | √ | Good |
| Weichenthal et al. 2017 | √ | √ | √ | √ | √ | √ | √ | √ | √ | √ | √ | √ | √ | √ | Good |
| Cakmak et al. 2018 | √ | √ | √ | √ | √ | √ | √ | √ | √ | √ | √ | √ | √ | √ | Good |
| Di et al. 2018 | √ | √ | √ | | √ | √ | √ | √ | √ | √ | √ | √ | √ | √ | Fair |
| Hvidtfeldt et al. 2019 | √ | √ | √ | | √ | √ | | √ | √ | √ | √ | √ | √ | √ | Fair |
| Lim et al. 2019 | √ | √ | √ | √ | √ | √ | √ | √ | √ | √ | √ | √ | √ | √ | Good |
| Kazemiparkouhi et al. 2019 | √ | √ | √ | | √ | √ | √ | √ | | √ | √ | √ | √ | √ | Fair |
| Paul et al. 2020 | √ | √ | √ | √ | √ | √ | √ | √ | √ | √ | √ | √ | √ | √ | Good |

**Table S5** **GRADE assessment for epidemiological evidences on the association between long-term $O_3$ exposure and cause-specific mortality risks.**

**Mortality Cause: All Causes**

| **Domains** | **Assessment** | **Downgrade/Upgrade** |
|---|---|---|
| Start Level | All cohort studies. | High |
| Risk of Bias | The overall risk of bias in all cohorts was low. | No change |
| Imprecision | All studies included reported the 95% confidence interval around the best estimate of the absolute effect. | No change |
| Inconsistency | The values of effect sizes across the studies were inconsistent, as the point estimates were in the range of 0.79 to 1.09. | Downgrade |
| Indirectness | All studies included the desired population, exposures and outcomes. | No change |
| Publication Bias | Given the comprehensive search, it seemed little publication bias. | No change |
| Magnitude of Associations | The magnitude of effect sizes was not large enough to upgrade the level of evidence. | No change |
| Dose-Response Trend | Linear dose-response relationship was assumed in all studies, and Di et al. 2017 checked the dose-response trends. | Upgrade |
| Plausible Confounding towards Null | Cakmak et al. 2018 reported higher RR after adjusting confounders. | Upgrade |
| **Overall Judgment** | **High** | |

**Mortality Cause: All Respiratory Diseases**

| **Domains** | **Assessment** | **Downgrade/Upgrade** |
|---|---|---|
| Start Level | All cohort studies. | High |
| Risk of Bias | The overall risk of bias in all cohorts was low. | No change |
| Imprecision | All studies included reported the 95% confidence interval around the best estimate of the absolute effect. | No change |
| Inconsistency | The values of effect sizes across the studies were inconsistent, as the point estimates were in the range of 0.75 to 1.17 | Downgrade |
| Indirectness | All studies included the desired population, exposures and outcomes. | No change |
| Publication Bias | Given the comprehensive search, it seemed little publication bias. | No change |
| Magnitude of Associations | The magnitude of effect sizes was not large enough to upgrade the level of evidence. | No change |
| Dose-Response Trend | Linear dose-response relationship was assumed in all studies, and 2 out of 20 studies (Jerrett et al. 2009; Lim et al. 2019) tested dose-response trends. | Upgrade |
| Plausible Confounding towards Null | No crude and adjusted risks were provided for each study. | No change |
| **Overall Judgment** | **High** | |

**Mortality Cause: Chronic Obstructive Pulmonary Disease (COPD)**

| **Domains** | **Assessment** | **Downgrade/Upgrade** |
|---|---|---|
| Start Level | All cohort studies. | High |
| Risk of Bias | The overall risk of bias in all cohorts was low. | No change |
| Imprecision | All studies included reported the 95% confidence interval around the best estimate of the absolute effect. | No change |
| Inconsistency | The values of effect sizes across the studies were inconsistent, as the point estimates were in the range of 0.95 to 1.17. | Downgrade |
| Indirectness | All studies included the desired population, exposures and outcomes. | No change |
| Publication Bias | Given the comprehensive search, there is no need to consider publication bias. | No change |
| Magnitude of Associations | The magnitude of effect sizes was not large enough to upgrade the level of evidence. | No change |
| Dose-Response Trend | Linear dose-response relationship was assumed in all studies, but no studies considered dose-response trends. | No change |
| Plausible Confounding towards Null | No crude and adjusted risks were provided for each study. | No change |
| **Overall Judgment** | **Moderate** | |

**Table S5** (cont'd) **GRADE assessment for epidemiological evidences on the association between long-term $O_3$ exposure and cause-specific mortality risks.**

**Mortality Cause: Cardiovascular Diseases (CVD)**

| **Domains** | **Assessment** | **Downgrade/Upgrade** |
|---|---|---|
| Start Level | All cohort studies. | High |
| Risk of Bias | The overall risk of bias in all cohorts was low. | No change |
| Imprecision | All studies included reported the 95% confidence interval around the best estimate of the absolute effect. | No change |
| Inconsistency | The values of effect sizes across the studies were inconsistent, as the point estimates were in the range of 0.80 to 1.19. | Downgrade |
| Indirectness | All studies included the desired population, exposures and outcomes. | No change |
| Publication Bias | Positive publication bias was detected. | Downgrade |
| Magnitude of Associations | The magnitude of effect sizes was not large enough to upgrade the level of evidence. | No change |
| Dose-Response Trend | Linear dose-response relationship was assumed in all studies, and 2 studies (Lim et al. 2019; Paul et al. 2020) checked the dose-response trends. | Upgrade |
| Plausible Confounding towards Null | No crude and adjusted risks were provided for each study. | No change |
| **Overall Judgment** | **Moderate** | |

**Mortality Cause: Ischaemic Heart Disease (IHD)**

| **Domains** | **Assessment** | **Downgrade/Upgrade** |
|---|---|---|
| Start Level | All cohort studies. | High |
| Risk of Bias | The overall risk of bias in all cohorts was low. | No change |
| Imprecision | All studies included reported the 95% confidence interval around the best estimate of the absolute effect. | No change |
| Inconsistency | The values of effect sizes across the studies were inconsistent, as the point estimates were in the range of 0.96 to 1.14. | Downgrade |
| Indirectness | All studies included the desired population, exposures and outcomes. | No change |
| Publication Bias | Given the comprehensive search, there is no need to consider publication bias. | No change |
| Magnitude of Associations | The magnitude of effect sizes was not large enough to upgrade the level of evidence. | No change |
| Dose-Response Trend | Linear dose-response relationship was assumed in all studies, but no studies considered dose-response trends. | No change |
| Plausible Confounding towards Null | Cakmak et al. 2018 reported higher RR after adjusting confounders. | No change |
| **Overall Judgment** | **Moderate** | |

**Mortality Cause: Lung Cancer (LC)**

| **Domains** | **Assessment** | **Downgrade/Upgrade** |
|---|---|---|
| Start Level | All cohort studies. | High |
| Risk of Bias | The overall risk of bias in all cohorts was low. | No change |
| Imprecision | All studies included reported the 95% confidence interval around the best estimate of the absolute effect. | No change |
| Inconsistency | The values of effect sizes across the studies were inconsistent, as the point estimates were in the range of 0.80 to 1.85. | Downgrade |
| Indirectness | All studies included the desired population, exposures and outcomes. | No change |
| Publication Bias | Given the comprehensive search, it seemed little publication bias. | No change |
| Magnitude of Associations | The magnitude of effect sizes was not large enough to upgrade the level of evidence. | No change |
| Dose-Response Trend | Linear dose-response relationship was assumed in all studies, but no studies considered dose-response trends. | No change |
| Plausible Confounding towards Null | Cakmak et al. 2018 reported higher RR after adjusting confounders. | Upgrade |
| **Overall Judgment** | **High** | |

**Table S5** (cont'd) **GRADE assessment for epidemiological evidences on the association between long-term $O_3$ exposure and cause-specific mortality risks.**

| **Mortality Cause: Cerebrovascular Disease (CEVD)** | | |
|---|---|---|
| **Domains** | **Assessment** | **Downgrade/Upgrade** |
| Start Level | All cohort studies. | High |
| Risk of Bias | The overall risk of bias in all cohorts was low. | No change |
| Imprecision | All studies included reported the 95% confidence interval around the best estimate of the absolute effect. | No change |
| Inconsistency | The values of effect sizes across the studies were inconsistent, as the point estimates were in the range of 0.91 to 1.03. | Downgrade |
| Indirectness | All studies included the desired population, exposures and outcomes. | No change |
| Publication Bias | Positive publication bias was detected. | Downgrade |
| Magnitude of Associations | The magnitude of effect sizes was not large enough to upgrade the level of evidence. | No change |
| Dose-Response Trend | Linear dose-response relationship was assumed in all studies, but no studies considered dose-response trends. | No change |
| Plausible Confounding towards Null | No crude and adjusted risks were provided for each study. | No change |
| **Overall Judgment** | **Low** | |

| **Mortality Cause: Congestive Heart Failure (CHF)** | | |
|---|---|---|
| **Domains** | **Assessment** | **Downgrade/Upgrade** |
| Start Level | All cohort studies. | High |
| Risk of Bias | The overall risk of bias in all cohorts was low. | No change |
| Imprecision | All studies included reported the 95% confidence interval around the best estimate of the absolute effect. | No change |
| Inconsistency | The values of effect sizes across the studies were consistent, as the point estimates were in the range of 1.01 to 1.30. | Upgrade |
| Indirectness | All studies included the desired population, exposures and outcomes | No change |
| Publication Bias | Given the comprehensive search, it seemed little publication bias. | No change |
| Magnitude of Associations | The magnitude of effect sizes was not large enough to upgrade the level of evidence. | No change |
| Dose-Response Trend | Linear dose-response relationship was assumed in all studies, but no studies considered dose-response trends. | No change |
| Plausible Confounding towards Null | No crude and adjusted risks were provided for each study. | No change |
| **Overall Judgment** | **High** | |

**Table S6** **One-dropout sensitivity analysis of long-term $O_3$ exposure-associated all-cause, respiratory diseases, COPD, cardiovascular diseases, congestive heart failure, ischaemic heart disease, cerebrovascular diseases, and lung cancer mortality risks by 6mDMA8 exposure metric.** The pooled relative risks (RR), 95% confidence intervals (95% CI) presented in lower bounds (LB) and upper bounds (UB), and heterogeneity $I^2$ were estimated after excluding each study.

| Study | RR | LB | UB | $I^2$ |
|---|---|---|---|---|
| **All-cause Mortality** | | | | |
| Abbey et al. 1999 | 1.007 | 1.003 | 1.011 | 93.9% |
| Lipfert et al. 2006 | 1.006 | 1.002 | 1.010 | 93.5% |
| Jerrett et al. 2009 | 1.009 | 1.004 | 1.013 | 93.2% |
| Krewski et al. 2009 | 1.006 | 1.002 | 1.010 | 93.4% |
| Smith et al. 2009 | 1.008 | 1.003 | 1.012 | 93.6% |
| Lipsett et al. 2011 | 1.008 | 1.003 | 1.013 | 93.4% |
| Carey et al. 2013 | 1.008 | 1.003 | 1.013 | 93.3% |
| Jerrett et al. 2013 | 1.008 | 1.003 | 1.012 | 93.6% |
| Bentayeb et al. 2015 | 1.007 | 1.003 | 1.012 | 93.5% |
| Crouse et al. 2015 | 1.006 | 1.002 | 1.010 | 93.4% |
| Tonne et al. 2016 | 1.007 | 1.003 | 1.012 | 93.6% |
| Turner et al. 2016 | 1.006 | 1.002 | 1.011 | 93.5% |
| Weichenthal et al. 2017 | 1.003 | 0.999 | 1.007 | 86.9% |
| Cakmak et al. 2018 | 1.007 | 1.002 | 1.011 | 93.5% |
| Di et al. 2018 | 1.007 | 0.997 | 1.016 | 92.9% |
| Hvidtfeldt et al. 2019 | 1.009 | 1.003 | 1.013 | 93.4% |
| Kazemiparkouhi et al. 2019 | 1.007 | 0.998 | 1.016 | 92.6% |
| Lim et al. 2019 | 1.008 | 1.003 | 1.012 | 93.6% |
| **Respiratory Mortality** | | | | |
| Abbey et al. 1999 | 1.020 | 1.000 | 1.041 | 89.4% |
| Jerrett et al. 2009 | 1.018 | 0.997 | 1.041 | 88.4% |
| Smith et al. 2009 | 1.020 | 0.997 | 1.042 | 88.5% |
| Lipsett et al. 2011 | 1.021 | 0.999 | 1.043 | 88.3% |
| Carey et al. 2013 | 1.028 | 1.012 | 1.046 | 78.8% |
| Jerrett et al. 2013 | 1.022 | 1.001 | 1.045 | 87.9% |
| Bentayeb et al. 2015 | 1.022 | 1.001 | 1.042 | 88.5% |
| Crouse et al. 2015 | 1.025 | 1.004 | 1.046 | 86.7% |
| Turner et al. 2016 | 1.014 | 0.994 | 1.034 | 86.3% |
| Weichenthal et al. 2017 | 1.019 | 0.997 | 1.041 | 88.5% |
| Hvidtfeldt et al. 2019 | 1.024 | 1.003 | 1.046 | 86.7% |
| Kazemiparkouhi et al. 2019 | 1.019 | 0.994 | 1.045 | 85.2% |
| Lim et al. 2019 | 1.019 | 0.998 | 1.041 | 88.5% |
| **COPD Mortality** | | | | |
| Zanobetti et al. 2011 | 1.047 | 0.984 | 1.112 | 95.6% |
| Crouse et al. 2015 | 1.083 | 1.035 | 1.134 | 91.0% |
| Turner et al. 2016 | 1.051 | 0.988 | 1.118 | 95.7% |
| Cakmak et al. 2018 | 1.075 | 1.016 | 1.137 | 93.1% |
| Kazemiparkouhi et al. 2019 | 1.053 | 0.986 | 1.124 | 91.9% |
| Lim et al. 2019 | 1.062 | 0.998 | 1.129 | 95.7% |
| **Cardiovascular Mortality** | | | | |
| Jerrett et al. 2009 | 1.031 | 1.003 | 1.059 | 98.1% |
| Smith et al. 2009 | 1.028 | 0.999 | 1.057 | 98.1% |
| Lipsett et al. 2011 | 1.028 | 1.000 | 1.058 | 98.1% |
| Jerrett et al. 2013 | 1.028 | 1.000 | 1.058 | 98.1% |
| Bentayeb et al. 2015 | 1.028 | 1.003 | 1.053 | 98.1% |
| Crouse et al. 2015 | 1.025 | 0.998 | 1.053 | 98.0% |
| Turner et al. 2016 | 1.027 | 0.999 | 1.054 | 98.1% |
| Weichenthal et al. 2017 | 1.014 | 0.999 | 1.028 | 92.8% |
| Hvidtfeldt et al. 2019 | 1.033 | 1.005 | 1.060 | 98.1% |
| Kazemiparkouhi et al. 2019 | 1.029 | 0.997 | 1.063 | 97.5% |
| Lim et al. 2019 | 1.027 | 1.000 | 1.055 | 98.1% |
| Paul et al. 2020 | 1.020 | 0.995 | 1.046 | 97.9% |
| **Congestive Heart Failure Mortality** | | | | |
| Zanobetti et al. 2011 | 1.076 | 1.025 | 1.129 | 88.2% |
| Turner et al. 2016 | 1.058 | 1.020 | 1.096 | 86.6% |
| Kazemiparkouhi et al. 2019 | 1.065 | 1.009 | 1.126 | 87.5% |
| Lim et al. 2019 | 1.089 | 1.061 | 1.116 | 73.4% |
| **Ischaemic Heart Disease Mortality** | | | | |
| Jerrett et al. 2009 | 1.030 | 0.997 | 1.065 | 97.9% |
| Krewski et al. 2009 | 1.024 | 0.990 | 1.060 | 98.0% |
| Lipsett et al. 2011 | 1.023 | 0.989 | 1.059 | 98.0% |
| Jerrett et al. 2013 | 1.023 | 0.989 | 1.059 | 98.0% |
| Crouse et al. 2015 | 1.018 | 0.985 | 1.052 | 97.7% |
| Turner et al. 2016 | 1.028 | 0.994 | 1.063 | 98.0% |
| Cakmak et al. 2018 | 1.011 | 0.992 | 1.030 | 92.1% |
| Kazemiparkouhi et al. 2019 | 1.027 | 0.990 | 1.065 | 96.9% |
| Lim et al. 2019 | 1.022 | 0.989 | 1.057 | 98.0% |
| **Cerebrovascular Mortality** | | | | |
| Lipsett et al. 2011 | 0.995 | 0.963 | 1.028 | 79.1% |
| Crouse et al. 2015 | 0.990 | 0.966 | 1.014 | 68.9% |
| Turner et al. 2016 | 0.990 | 0.966 | 1.015 | 72.7% |
| Kazemiparkouhi et al. 2019 | 1.000 | 0.967 | 1.034 | 69.5% |
| Lim et al. 2019 | 1.003 | 0.982 | 1.025 | 72.2% |
| **Lung Cancer Mortality** | | | | |
| Abbey et al. 1999 | 0.984 | 0.962 | 1.006 | 84.2% |
| Krewski et al. 2009 | 0.984 | 0.959 | 1.009 | 83.1% |
| Lipsett et al. 2011 | 0.984 | 0.959 | 1.009 | 83.0% |
| Carey et al. 2013 | 0.989 | 0.967 | 1.011 | 79.1% |
| Jerrett et al. 2013 | 0.986 | 0.963 | 1.010 | 80.9% |
| Crouse et al. 2015 | 0.986 | 0.963 | 1.010 | 80.1% |
| Turner et al. 2016 | 0.986 | 0.963 | 1.010 | 81.9% |
| Cakmak et al. 2018 | 0.981 | 0.957 | 1.004 | 83.4% |
| Kazemiparkouhi et al. 2019 | 0.976 | 0.963 | 0.990 | 23.0% |
| Lim et al. 2019 | 0.984 | 0.960 | 1.010 | 81.7% |

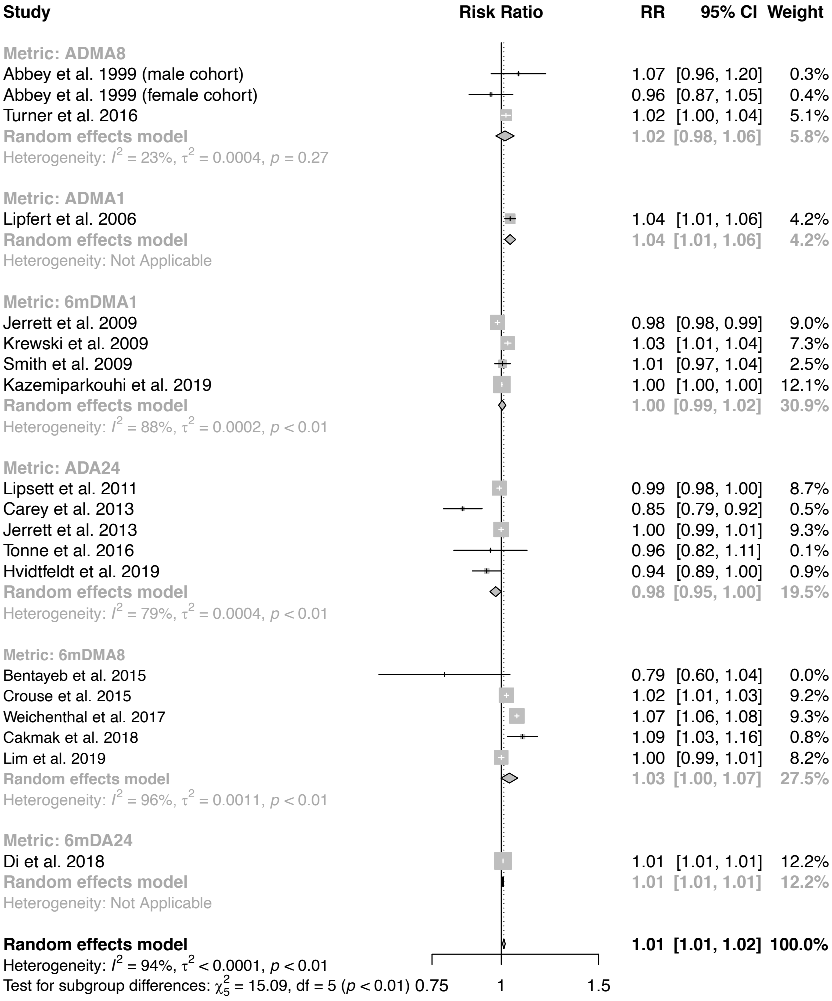

**Figure S1** Sub-group meta-analyses by metrics of non-accidental all-cause mortality risks attributable to every 10-ppbV incremental long-term $O_3$ exposure.

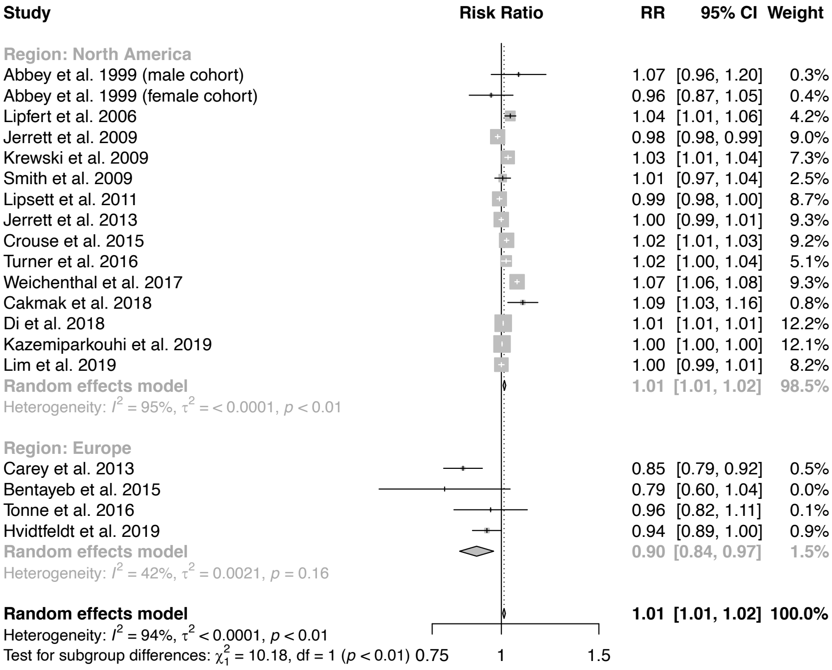

**Figure S2** Sub-group meta-analyses by cohort population ethnicity of non-accidental all-cause mortality risks attributable to every 10-ppbV incremental long-term $O_3$ exposure.

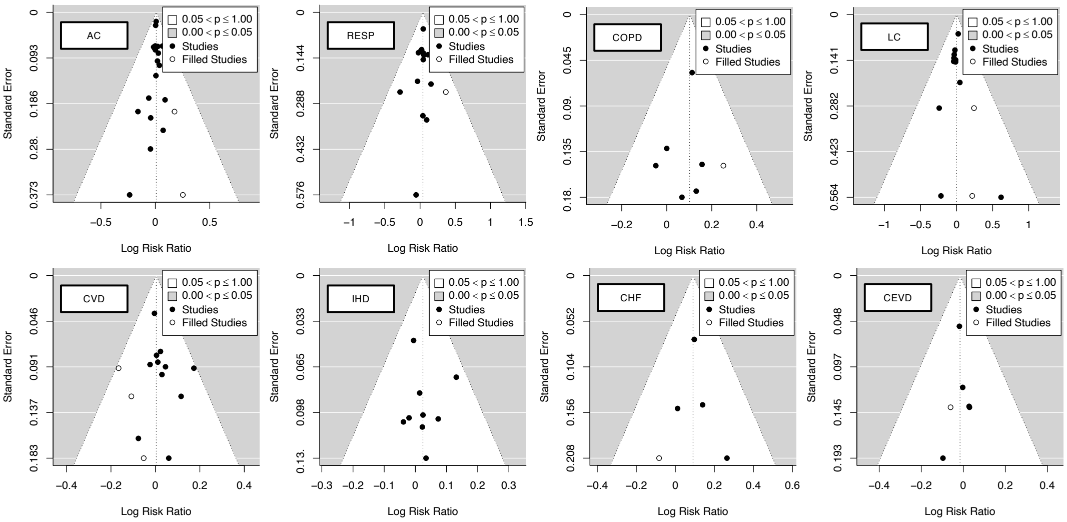

**Figure S3 Publication bias test by trim-and-fill method for 9 causes of $O_3$-exposure attributable mortality risks.** Biases were highlighted by statistically filled values to reach symmetry between risks higher and lower than pooled values.

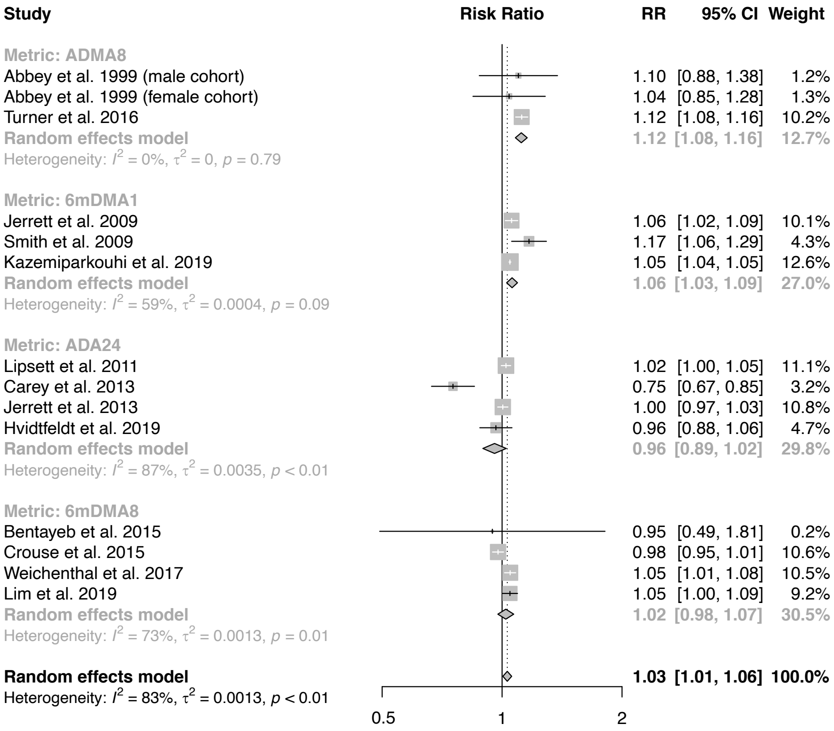

**Figure S4 Sub-group meta-analyses by metrics of all respiratory mortality risks attributable to every 10-ppbV incremental long-term $O_3$ exposure.**

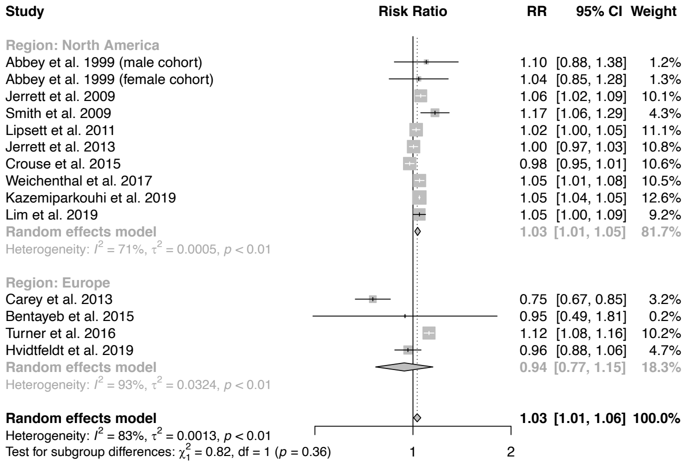

**Figure S5 Sub-group meta-analyses by cohort population ethnicity of all respiratory mortality risks attributable to every 10-ppbV incremental long-term $O_3$ exposure.**

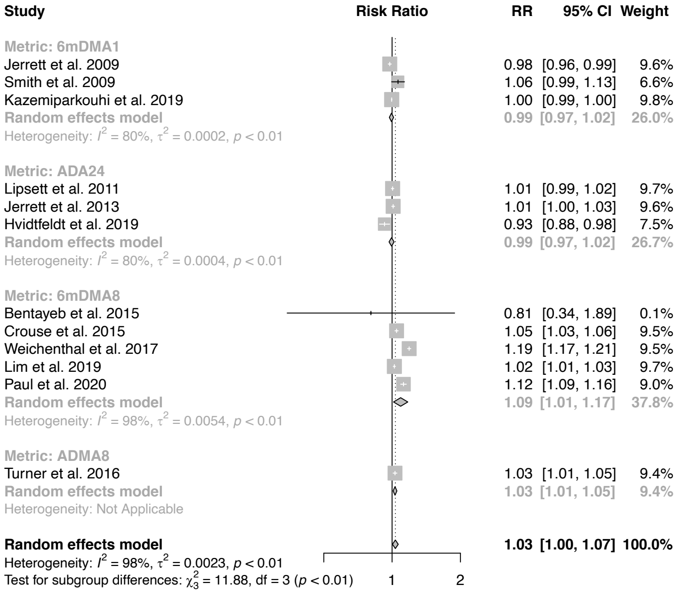

**Figure S6 Sub-group meta-analyses by metrics of cardiovascular mortality risks attributable to every 10-ppbV incremental long-term $O_3$ exposure.**

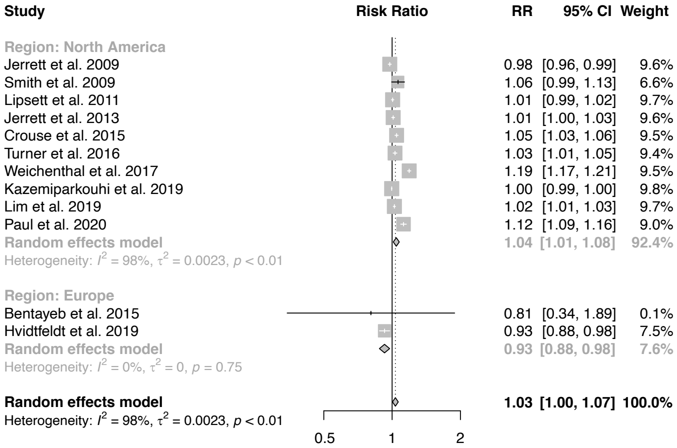

**Figure S7 Sub-group meta-analyses by cohort population ethnicity of cardiovascular mortality risks attributable to every 10-ppbV incremental long-term $O_3$ exposure.**

### REFERENCES

1. Bell, M. L.; Peng, R. D.; Dominici, F., The exposure-response curve for ozone and risk of mortality and the adequacy of current ozone regulations. *Environ Health Perspect* **2006,** *114*, (4), 532-6.
2. Burnett, R. T.; Pope, C. A., 3rd; Ezzati, M.; Olives, C.; Lim, S. S.; Mehta, S.; Shin, H. H.; Singh, G.; Hubbell, B.; Brauer, M.; Anderson, H. R.; Smith, K. R.; Balmes, J. R.; Bruce, N. G.; Kan, H.; Laden, F.; Pruss-Ustun, A.; Turner, M. C.; Gapstur, S. M.; Diver, W. R.; Cohen, A., An integrated risk function for estimating the global burden of disease attributable to ambient fine particulate matter exposure. *Environ Health Perspect* **2014,** *122*, (4), 397-403.
3. Malley, C. S.; Henze, D. K.; Kuylenstierna, J. C. I.; Vallack, H. W.; Davila, Y.; Anenberg, S. C.; Turner, M. C.; Ashmore, M. R., Updated Global Estimates of Respiratory Mortality in Adults ≥30 Years of Age Attributable to Long-Term Ozone Exposure. *Environ Health Perspect* **2017,** *125*, (8), 087021.
4. Lipfert, F. W.; Wyzga, R. E.; Baty, J. D.; Miller, J. P., Traffic density as a surrogate measure of environmental exposures in studies of air pollution health effects: Long-term mortality in a cohort of US veterans. *Atmos Environ* **2006,** *40*, (1), 154-169.
5. Smith, K. R.; Jerrett, M.; Anderson, H. R.; Burnett, R. T.; Stone, V.; Derwent, R.; Atkinson, R. W.; Cohen, A.; Shonkoff, S. B.; Krewski, D.; Pope, C. A., 3rd; Thun, M. J.; Thurston, G., Public health benefits of strategies to reduce greenhouse-gas emissions: health implications of short-lived greenhouse pollutants. *Lancet* **2009,** *374*, (9707), 2091-2103.
6. Lipsett, M. J.; Ostro, B. D.; Reynolds, P.; Goldberg, D.; Hertz, A.; Jerrett, M.; Smith, D. F.; Garcia, C.; Chang, E. T.; Bernstein, L., Long-term exposure to air pollution and cardiorespiratory disease in the California teachers study cohort. *Am J Respir Crit Care Med* **2011,** *184*, (7), 828-35.
7. Carey, I. M.; Atkinson, R. W.; Kent, A. J.; van Staa, T.; Cook, D. G.; Anderson, H. R., Mortality associations with long-term exposure to outdoor air pollution in a national English cohort. *Am J Respir Crit Care Med* **2013,** *187*, (11), 1226-33.
8. Bentayeb, M.; Wagner, V.; Stempfelet, M.; Zins, M.; Goldberg, M.; Pascal, M.; Larrieu, S.; Beaudeau, P.; Cassadou, S.; Eilstein, D., Association between long-term exposure to air pollution and mortality in France: a 25-year follow-up study. *Environ Int* **2015,** *85*, 5-14.
9. Crouse, D. L.; Peters, P. A.; Hystad, P.; Brook, J. R.; van Donkelaar, A.; Martin, R. V.; Villeneuve, P. J.; Jerrett, M.; Goldberg, M. S.; Pope III, C. A., Ambient $PM_{2.5}$, $O_3$, and $NO_2$ exposures and associations with mortality over 16 years of follow-up in the Canadian Census Health and Environment Cohort (CanCHEC). *Environ Health Perspect* **2015,** *123*, (11), 1180-1186.
10. Di, Q.; Wang, Y.; Zanobetti, A.; Wang, Y.; Koutrakis, P.; Choirat, C.; Dominici, F.; Schwartz, J. D., Air Pollution and Mortality in the Medicare Population. *N Engl J Med* **2017,** *376*, (26), 2513-2522.
11. Hvidtfeldt, U. A.; Sorensen, M.; Geels, C.; Ketzel, M.; Khan, J.; Tjonneland, A.; Overvad, K.; Brandt, J.; Raaschou-Nielsen, O., Long-term residential exposure to $PM_{2.5}$, $PM_{10}$, black carbon, $NO_2$, and ozone and mortality in a Danish cohort. *Environ Int* **2019,** *123*, 265-272.
12. Kazemiparkouhi, F.; Eum, K. D.; Wang, B.; Manjourides, J.; Suh, H. H., Long-term ozone exposures and cause-specific mortality in a US Medicare cohort. *J Exp Sci Environ Epidemiol* **2020,** *30*, (4), 650-658.
13. Krewski, D.; Jerrett, M.; Burnett, R. T.; Ma, R.; Hughes, E.; Shi, Y.; Turner, M. C.; Pope III, C. A.; Thurston, G.; Calle, E. E., *Extended follow-up and spatial analysis of the American Cancer Society study linking particulate air pollution and mortality*. Health Effects Institute Boston, MA: 2009.
14. Abbey, D. E.; Nishino, N.; McDonnell, W. F.; Burchette, R. J.; Knutsen, S. F.; Lawrence Beeson, W.; Yang, J. X., Long-term inhalable particles and other air pollutants related to mortality in nonsmokers. *Am J Respir Crit Care Med* **1999,** *159*, (2), 373-82.
15. Jerrett, M.; Burnett, R. T.; Pope, C. A., 3rd; Ito, K.; Thurston, G.; Krewski, D.; Shi, Y.; Calle, E.; Thun, M., Long-term ozone exposure and mortality. *N Engl J Med* **2009,** *360*, (11), 1085-95.
16. Jerrett, M.; Burnett, R. T.; Beckerman, B. S.; Turner, M. C.; Krewski, D.; Thurston, G.; Martin, R. V.; van Donkelaar, A.; Hughes, E.; Shi, Y.; Gapstur, S. M.; Thun, M. J.; Pope, C. A., 3rd, Spatial analysis of air pollution and mortality in California. *Am J Respir Crit Care Med* **2013,** *188*, (5), 593-9.
17. Zanobetti, A.; Schwartz, J., Ozone and survival in four cohorts with potentially predisposing diseases. *Am J Respir Crit Care Med* **2011,** *184*, (7), 836-41.
18. Lim, C. C.; Hayes, R. B.; Ahn, J.; Shao, Y.; Silverman, D. T.; Jones, R. R.; Garcia, C.; Bell, M. L.; Thurston, G. D., Long-Term Exposure to Ozone and Cause-Specific Mortality Risk in the United States. *Am J Respir Crit Care Med* **2019,** *200*, (8), 1022-1031.
19. Paul, L. A.; Burnett, R. T.; Kwong, J. C.; Hystad, P.; van Donkelaar, A.; Bai, L.; Goldberg, M. S.; Lavigne, E.; Copes, R.; Martin, R. V., The impact of air pollution on the incidence of diabetes and survival among prevalent diabetes cases. *Environ Int* **2020,** *134*, 105333.
20. Cakmak, S.; Hebbern, C.; Pinault, L.; Lavigne, E.; Vanos, J.; Crouse, D. L.; Tjepkema, M., Associations between long-term $PM_{2.5}$ and ozone exposure and mortality in the Canadian Census Health and Environment Cohort (CANCHEC), by spatial synoptic classification zone. *Environ Int* **2018,** *111*, 200-211.
